# A phenome-wide association study (PheWAS) of COVID-19 outcomes by race using the electronic health records data in Michigan Medicine

**DOI:** 10.1101/2020.06.29.20141564

**Authors:** Maxwell Salvatore, Tian Gu, Jasmine A. Mack, Swaraaj Prabhu Sankar, Snehal Patil, Thomas S. Valley, Karandeep Singh, Brahmajee K. Nallamothu, Sachin Kheterpal, Lynda Lisabeth, Lars G. Fritsche, Bhramar Mukherjee

**Author notes:** Corresponding author: Bhramar Mukherjee, University of Michigan School of Public Health Department of Biostatistics, 1415 Washington Heights, Ann Arbor, MI 48109, United States.

## Abstract

**Background:** We perform a phenome-wide scan to identify pre-existing conditions related to COVID-19 susceptibility and prognosis across the medical phenome and how they vary by race.

**Methods:** The study is comprised of 53,853 patients who were tested/positive for COVID-19 between March 10 and September 2, 2020 at a large academic medical center.

**Results:** Pre-existing conditions strongly associated with <u>hospitalization</u> were *renal failure, pulmonary heart disease*, and *respiratory failure*. Hematopoietic conditions were associated with <u>ICU admission/mortality</u> and mental disorders were associated with <u>mortality</u> in non-Hispanic Whites. Circulatory system and genitourinary conditions were associated with <u>ICU admission/mortality</u> in non-Hispanic Blacks.

**Conclusions:** Understanding pre-existing clinical diagnoses related to COVID-19 outcomes informs the need for targeted screening to support specific vulnerable populations to improve disease prevention and healthcare delivery.

## 1. Introduction

The emergence of electronic health records (EHR) and rise of EHR-linked biobanks has made it possible for researchers to explore -omics-based relationships agnostically on a large scale instead of targeted hypothesis testing. Introduced by Denny et al. in 2010, a phenome-wide association study (PheWAS) is an omnibus scan to identify gene-disease associations across the medical phenome[1]. Due to computational advances and development of widely available analytic frameworks[2–6], PheWAS are now relatively easy to implement. The main goal of a PheWAS is to replicate known gene-disease relationships and to search for hidden and unanticipated associations.

As of January 15, 2021, there are 23,759,743 confirmed COVID-19 cases in the US,[7] representing approximately 25% of all global cases. Because COVID-19 is a respiratory disease and produces flu-like symptoms, testing strategies in the US initially focused on those with symptoms, the elderly, and those with pre-existing conditions [8] - populations who are at risk of severe disease and complications. However, because COVID-19 is a novel disease, only a handful of pre-existing phenotypes are known to be associated with developing symptoms or experiencing adverse outcomes. These include liver, kidney, heart, and respiratory disease.

There has been a remarkable surge within the academic and medical communities to conduct rapid research on COVID-19[9]. However, only recently have there been studies examining disparities across an ensemble of COVID-19 associated conditions and outcomes in US patient cohorts[10–17]. Instead of a hypothesis driven approach based on the literature, this study applies an agnostic *disease-disease* PheWAS framework to COVID-19 outcomes in a cohort of 53,853 patients who were tested for or treated at a large academic medical center. We look at prognosis among all COVID-19 patients as well as separately among non-Hispanic White (White) and non-Hispanic Black/African American (Black) patients. The primary objective of this study is to agnostically identify conditions present in an individual’s medical record that may be associated with hospitalization, ICU admission, and mortality. We also present the results from race stratified susceptibility PheWAS in the supplementary materials. Our reason to downplay the outcome of who gets COVID-19 or who tests positive for COVID-19 is due to the prioritized testing strategy that makes it hard to find a suitable comparison group. A naïve comparison of the positive versus negative test results is highly biased.[17] However, conditional on testing positive, downstream outcomes are less prone to such selection biases and we focus on these outcomes.

## 2. Materials and Methods

### 2.1. Study design

#### 2.1.1. COVID-19 cohort

We extracted the EHR for patients tested for COVID-19 at the University of Michigan Health System, also known as Michigan Medicine (MM), from March 10, 2020 to September 2, 2020. A total of 53,260 patients (98.9%) who were tested at MM and 593 patients (1.1%) who were treated for COVID-19 in MM, but tested elsewhere, constituted our initial study cohort of 53,853 patients, of whom 2,582 tested positive. Since the testing protocol in MM[18] focused on prioritized testing, this is a non-random sample of the population. Study protocols were reviewed and approved by the University of Michigan Medical School Institutional Review Board (IRB ID HUM00180294).

### 2.2. Data Processing

#### 2.2.1. Classifying patients who were still in hospital and ICU

We categorized patients into non-hospitalized, hospitalized (includes ICU stays), and hospitalized with ICU stay based on the admission and discharge data. A total of 22 patients were still admitted in the hospital (17 had at least one ICU state and five had no ICU stay).

#### 2.2.2. Generation of the medical phenome

We constructed the medical phenome by extracting available International Classification of Diseases (ICD; ninth and tenth editions) code from EHR and forming them up to 1,813 traits using the PheWAS R package (as described in detail elsewhere).[1] Each of these traits (PheWAS codes) was coded as a binary risk factor (present/absent) and used as a predictor in the association models with COVID-19 outcomes. To differentiate *pre-existing* conditions from phenotypes related to COVID-19 testing/treatment, we applied a 14-day-prior restriction on the tested cohort by removing diagnoses that first appeared within the 14 days before the first test or diagnosis date, whichever was earlier. The analyses in this study were restricted to 1,363 traits that appeared in the EHR 14-day-prior of at least ten COVID-19 positive patients. Further, we realize that the aggregation of ICD codes into phecodes may result in clinically unusable or unclear phenotypes. While the PheWAS is performed on PheWAS codes, one can view the mapping of ICD-to-PheWAS code relationships on this website: https://prsweb.sph.umich.edu:8443/phecodeData/searchPhecode.

#### 2.2.3. Description of variables

A summary data dictionary is available with the source and definition of each variable used in our analysis (**Table S1A** in Supplement).

### 2.3. Statistical analysis

We performed PheWAS to identify predictors of three COVID-19 prognostic outcomes in this study (detailed definition in **Table S1B** in the Supplement), among those who were diagnosed/tested positive, comparing:

i. those who were hospitalized with those who were not
ii. those who were admitted to ICU or died with those who were not
iii. those who died with those who did not (no untested controls were used, only considers tested positive cohort)

We also present results from the susceptibility PheWAS (those who were diagnosed with COVID-19 with those who were not tested at all [matched controls]) in the supplementary materials.

All COVID-19 outcomes of interest are binary; thus, logistic regression was our primary tool. All logistic regression models were of the following form:

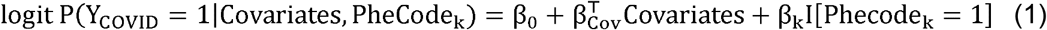

k = 1, …, 1051. Here Y_COVID_ is various COVID-19 related outcomes under consideration (e.g., COVID-19 hospitalization, ICU admission, and mortality). The Firth correction was used to address potential separation issues in logistic regression models.[19–21] Full models were adjusted for age, sex, race, and the neighborhood deprivation index (NDI). The NDI is defined by US census tract (corresponding to the residential address available in each patient’s EHR) for the year 2010 and are from the National Neighborhood Data Archive (NaNDA).[19] PheWAS adjusting for an additional comorbidity score covariate (indicating whether the patient was diagnosed with conditions across seven disease categories associated with COVID-19 susceptibility and adverse outcomes: respiratory, circulatory, any cancer, type II diabetes, kidney, liver, and autoimmune; ranges from 0 to 7) is included on our accompanying website: https://cphds.sph.umich.edu/covidphewas/.

#### 2.3.1. Race-stratified analysis

Since the prognostic factors could potentially be different across races, we carried out the entire analysis stratified by race. We restricted our attention to Whites and Blacks due to limitations of sample size for other racial groups. **Table S2** contains descriptive statistics stratified by race. We checked for the equality of the log(OR) corresponding to Whites and Blacks through a Wald test for the difference of the log(OR).

For all models, we report the Firth corrected estimate of the odds ratio, 95% Wald-type confidence interval and *P*-value. A conservative Bonferroni multiple testing correction was implemented to conclude statistically significant results (*P*=0.05/number of phecodes in analysis), and P < .05 was used as a threshold for suggestive traits.

## 3. Results

There were 53,853 patients who were either tested for or diagnosed with COVID-19 eligible for inclusion in this study. Of those eligible for inclusion, our study population comprised 47,862 individuals (n_tested_=47,862 [n_positive_=2,133]) who had available International Classification of Disease (ICD; ninth and tenth editions) code data after applying the 14-day-prior to testing restriction to the EHR. Furthermore, a total of 1,813 qualified ICD-code-based phenotypes, referred to as PheWAS codes, were initially screened of which 1,363 had at least 10 occurrences in our COVID-19 positive cohort and were included in the analysis.

Of those 53,853 who were tested for COVID-19, 44.2% (23,814) were males and the median age was 47 years. The majority were White (72.4% [38,977]) while 10.7% were Black (5,763). Out of the study cohort, 4.8% (2,582) were tested positive (**Table 1**).

**Table 1.**
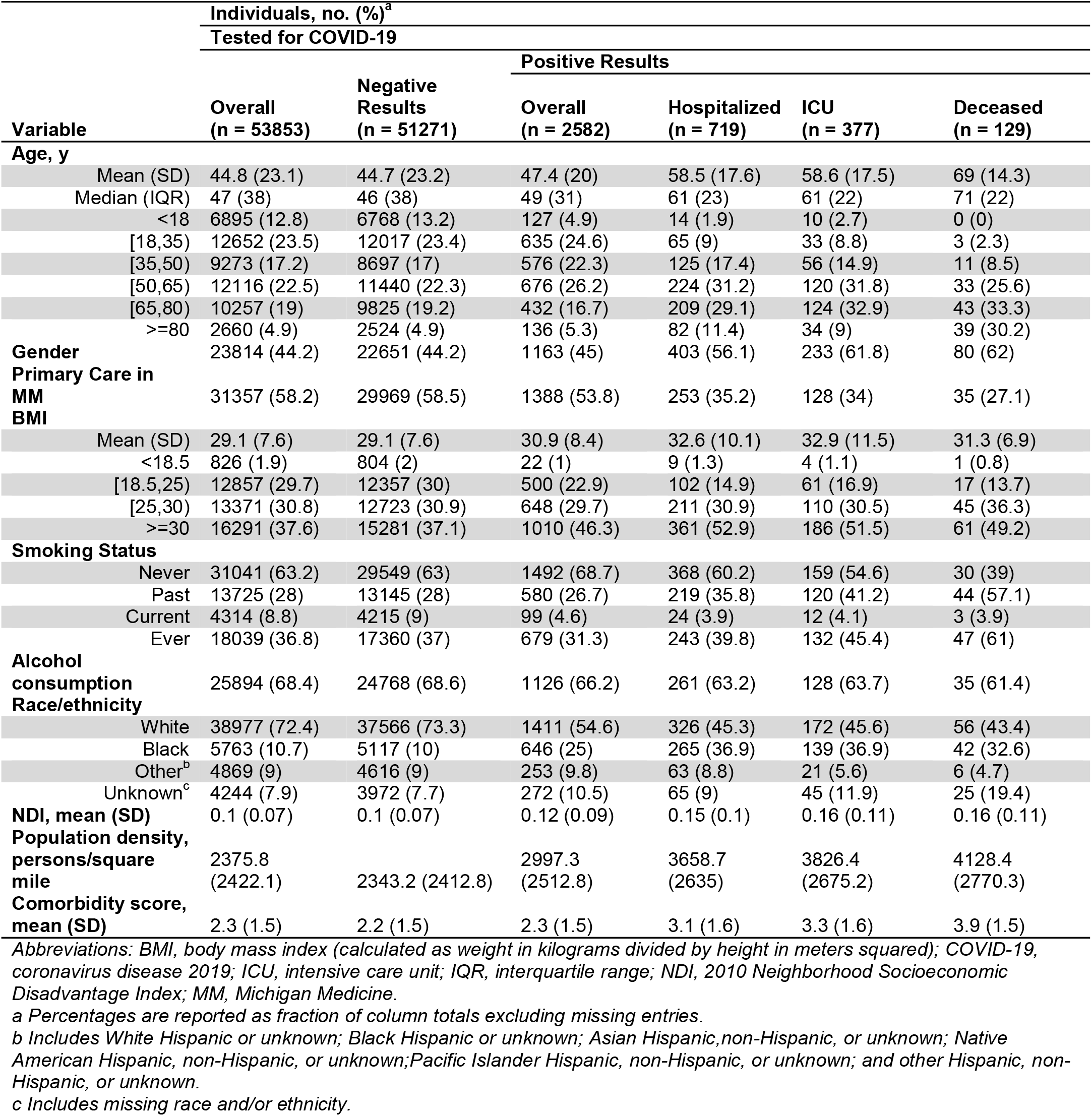
Descriptive Characteristics of the COVID-19 Tested/Diagnosed cohort at Michigan Medicine (March 10-September 2)

Among the 2,582 positive patients, 54.6% (1,411) were White, 25.0% (646) were Black, 27.8% (719) were hospitalized, 14.6% (377) were admitted to ICU and 5.0% (129) died.

### 3.1. Phenome-wide comorbidity association analysis

The association results for top 50 traits from the comorbidity PheWAS can be found in **Tables S3-S6** for the full cohort, Whites, and Blacks, side-by-side. Interactive versions of the PheWAS plots are online at https://cphds.sph.umich.edu/covidphewas/. This resource also provides tables with the adjusted odds ratios, 95% confidence intervals, p-values, and counts of occurrence in cases and controls for all traits included in the PheWAS performed.

#### 3.1.1. Full cohort prognostic associations

As the disease outcome progresses (from hospitalized to ICU, and to deceased), stronger associations with circulatory system, genitourinary (renal diseases in particular) and respiratory diseases were observed. Forty-four traits including 12 circulatory system and 11 respiratory diseases were phenome-wide significantly associated with hospitalization, as well as additional 263 suggestive traits under threshold of *P*<.05 (**Figure 1A**)— respiratory failure, insufficiency, arrest (P=3.98×10^−20^), acute renal failure (*P*=6.31×10^−13^), viral pneumonia (*P*=2.51×10^−11^), and acid-base balance disorder (*P*=2.40×10^−10^). Moreover, 58 phenome-wide significant hits (e.g., respiratory failure, insufficiency, arrest [*P*=1.58×10^−15^], acid-base balance disorder [*P*=3.98×10^−14^], and hypotension [*P*=1.58×10^−11^]) as well as 286 suggestive hits were noted for association with ICU admission/mortality (**Figure 1D**), including 77 circulatory system, 36 endocrine/metabolic, 35 genitourinary, and 31 respiratory diseases. There were 22 phenome-wide significant traits associated with COVID-19 mortality (**Figure 1G**), along with additional 227 suggestive traits under threshold *P*<0.05. In addition to 64 circulatory system and 31 endocrine/metabolic diseases, 23 mental disorders stood out as the third largest disease group associated with mortality, including delirium due to conditions classified elsewhere (*P*=9.33×10^−7^), memory loss (*P*=3.98×10^−4^) and aphasia (*P*=5.37×10^−4^).

**Figure 1.**
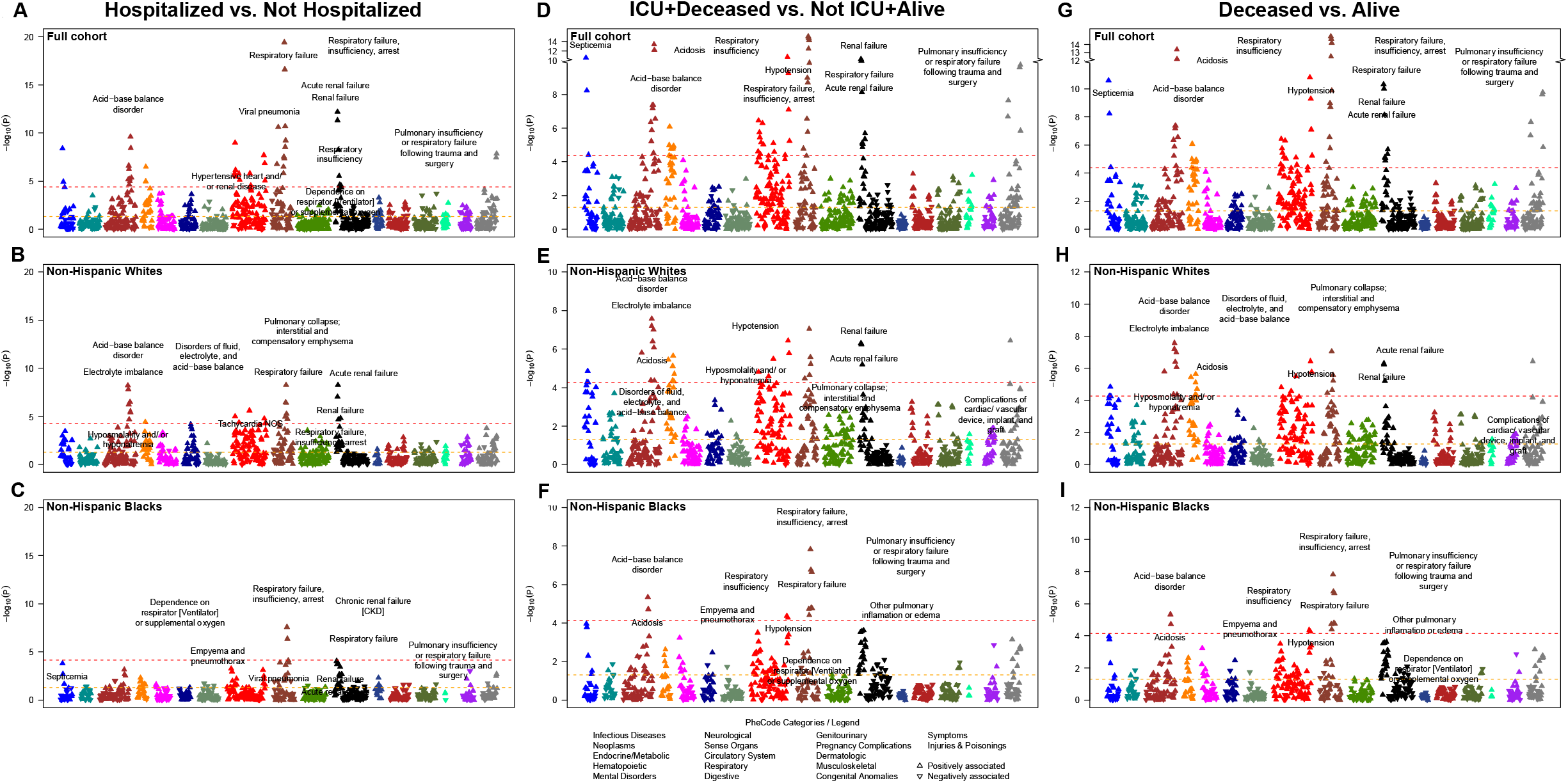
Manhattan plot showing the phenome-wide association between disease conditions and prognostic outcomes for COVID-19. Models are adjusted for age, sex, race (full cohort only), and three census tract-level socioeconomic indicators: proportion with less than high school education, proportion unemployed, and proportion with annual income below the federal poverty level. The x-axis are individual disease codes, color-coded by their corresponding disease category as described in the shared legend. The y-axis represents the -log_10_ transformed p-value of the association. The dashed, horizontal lines represent the p = 0.05 (in orange) and the Bonferroni corrected p-value (0.05 / number of tests; in red). Each point is represented by either an upward triangle indicating a positive association or a downward triangle indicating a negative association.

#### 3.1.2. Race-stratified prognostic associations

Among White patients, we identified 23 traits phenome-wide significantly associated with hospitalization (e.g., respiratory failure, insufficiency, arrest [*P*=5.25×10^−9^], acute renal failure [*P*=5.25×10^−9^], and electrolyte imbalance [*P*=1.51×10^−8^] **Figure 1B**), as well as 239 suggestive traits, including 54 circulatory system, 30 respiratory, 29 endocrine/metabolic, and 21 genitourinary diseases. Thirty-two phenome-wide significant traits (e.g., electrolyte imbalance [*P*=2.63×10^−8^], pulmonary collapse, interstitial and compensatory emphysema [*P*=8.91×10^−8^] and hypotension [*P*=3.63×10^−7^]) and 239 suggestive traits were associated with ICU admission/mortality (**Figure 1E**), including 60 circulatory system, 27 respiratory, 27 digestive, and 23 hematopoietic diseases. One phenome-wide significant trait (elevated white blood cell count [*P*=3.55×10^−5^]) and 130 suggestive traits were associated with COVID-19 mortality (**Figure 1H**), including 18 circulatory system, 17 endocrine/metabolic, 16 mental disorders, 14 genitourinary diseases such as osteomyelitis (*P*=1.74×10^−4^), neurological disorder (*P*=4.37×10^−4^) and aphasia (*P*=4.57×10^−4^).

Among Black patients, two phenome-wide significant traits were detected (respiratory failure, insufficiency, arrest [*P*=2.63×10^−8^], respiratory failure [*P*=4.37×10^−7^]) along with 89 traits nominally associated with hospitalization (**Figure 1C**), including 17 circulatory, 15 genitourinary, and 14 respiratory diseases. Eleven phenome-wide significant traits (e.g., respiratory failure, insufficiency, arrest [*P*=1.48×10^−8^], acid-base balance disorder [*P*=4.57×10^−6^], hypotension [*P*=4.37×10^−5^]) and 119 suggestive traits were associated with ICU admission/mortality, including 33 circulator, 26 genitourinary, and 17 endocrine/metabolic diseases. Six phenome-wide significant traits (e.g., empyema and pneumothorax [*P*=3.98×10^−5^], hyperosmolality and/or hypernatremia [*P*=5.37×10^−5^], atrial fibrillation [*P*=5.62×10^−5^]) and 105 suggestive traits were associated with mortality, including 34 circulator, 24 endocrine/metabolic, and 12 genitourinary diseases. As shown in **Figure 1H** and **Figure 1I**, the strength of association between circulatory system disorders and COVID-19 mortality was higher in Black patients compared with White. Similarly, we observe a higher prevalence of genitourinary diseases in Blacks associated with COVID-19 mortality such as stage I or II chronic kidney disease (*P*=2.34×10^−4^) compared with Whites.

When comparing the effect sizes of phenome-wide significant associations (in the full cohort) across Whites and Blacks, we found no significant differences in the effect sizes (though there are numerical differences) but these traits exhibited consistent risks among races for hospitalization (**Figure 1A**) and for ICU admission/mortality (**Figure 1B**).

#### 3.1.3. Summary Takeaways

In all cohorts, as the disease progressed to increasingly severe prognosis, the associated phenotypes concentrated in circulatory heart diseases and renal diseases (**Figure 2A**); pre-existing *cardiovascular system problems*, and chronic diseases such as *chronic pulmonary heart disease* and *chronic renal failure* appeared to be associated with poor prognosis, while mental disorders constituted the third largest category associated with COVID-19 mortality behind circulatory system and endocrine/metabolic diseases. When comparing the top 50 traits between Whites and Blacks, acidosis, pulmonary, acute/chronic renal diseases showed an association with hospitalization (**Figure 2B**) and ICU admission/mortality (**Figure 2C**) in both races, while acute renal consistently stood out as well as in mortality (**Figure 2D**). Effect estimates for parent phecodes and corresponding confidence intervals for phenome-wide significant traits by outcome by cohort are present in forest plots in **Figure 3**. A similar forest plot consisting of child phecodes are in **Figure S1**. A description of the susceptibility outcome results and corresponding PheWAS plots (**Figure S2**) are in the **supplementary materials**.

**Figure 2.**
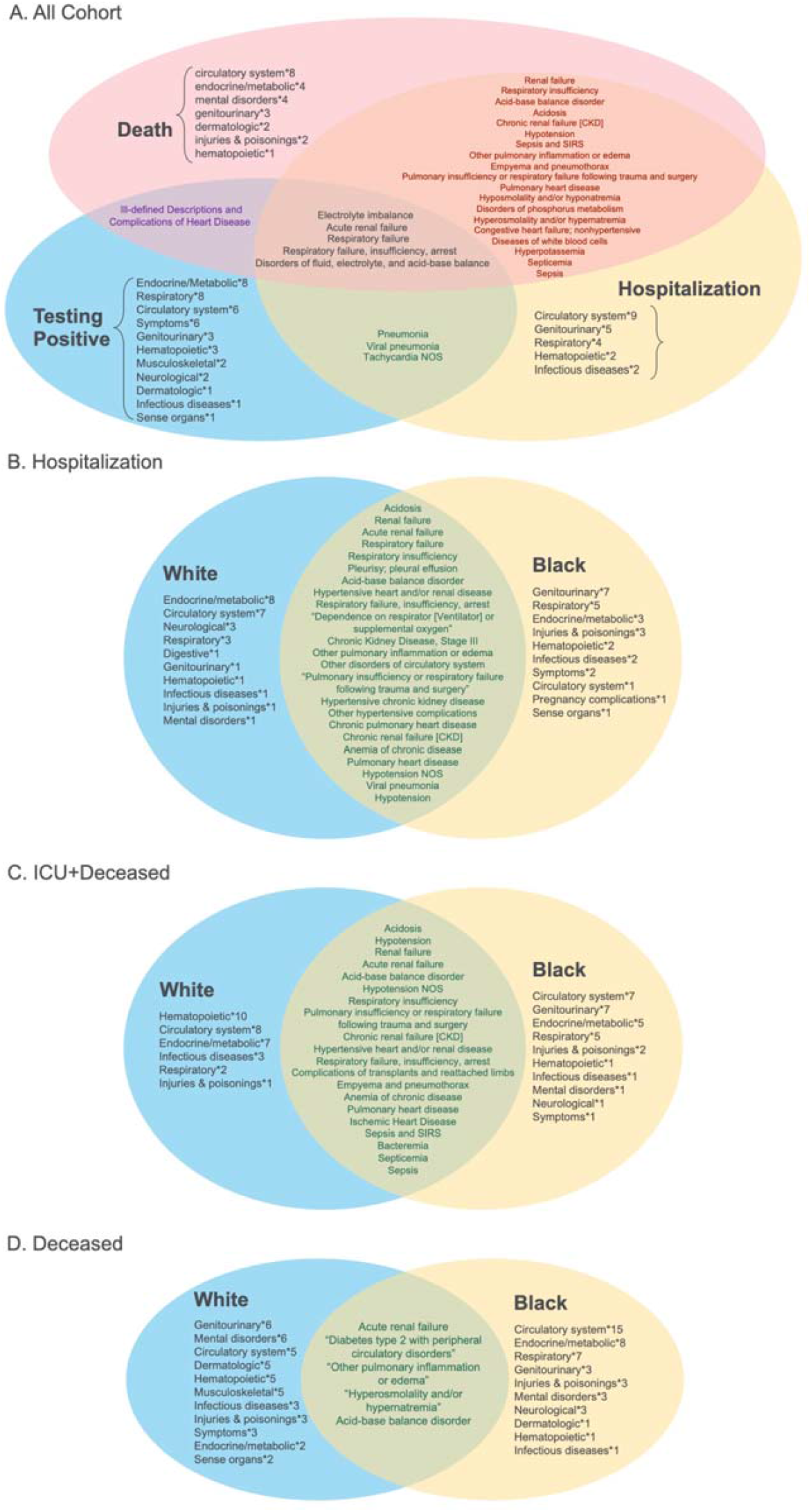
Venn diagrams of the top 50 traits. Each circle represents the top 50 hits from the full cohort PheWAS (panel A) and the racial PheWAS (panels B, C and D), respectively. Traits shared across PheWAS are stated, while the corresponding number of traits within a given disease category that are unique to that PheWAS are also provided.

**Figure 3.**
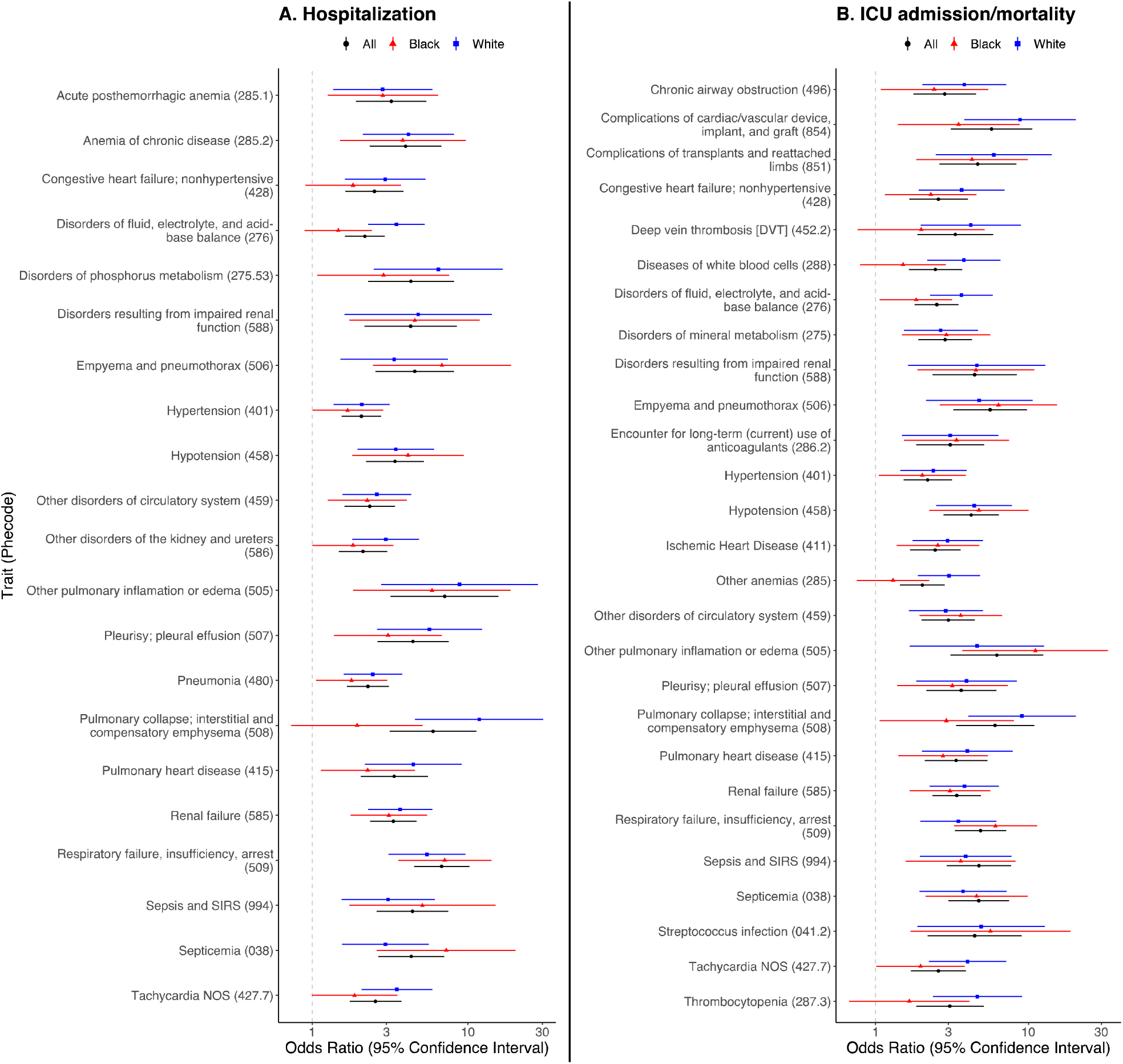
Forest plots of traits associated with poorer prognosis in overall cohort. A. Hospitalization. B. ICU admission/mortality. Odds ratios and 95% confidence intervals are shown for each trait whose PheWAS code is given in parentheses. Plots show parent codes only. Child codes found in Figure S

## 4. Discussion

Using data from a cohort of tested/diagnosed COVID-19 patients at MM, we performed what we believe is the first PheWAS looking at multiple COVID-19 outcomes stratified by race. Oetjens et al. recently published a PheWAS looking at phenotypes associated with COVID-19-related hospitalization, which is consistent with our results.[23] This technique allowed us to explore and identify potentially associated conditions across the medical phenome that are associated with susceptibility, hospitalization, ICU admission or mortality. Our results yield many previously known or plausibly associated phenotypes for increasingly severe prognosis such as pulmonary heart disease, respiratory failure and type 2 diabetes. Our stratified analysis showed that respiratory conditions and mental disorders appear to be associated with more severe outcomes among Whites while coagulation renal disease and heart disease are more strongly associated with severe outcomes among Blacks. **Figures 2B-2D** show that the disease categories that comprise the top 50 hits by prognostic outcome by race are different (with the caveat that for ICU admission and mortality these hits are largely suggestive due to limited power). Our results can inform targeted prevention across racial groups, which includes increased testing and encouraging self-isolation from household members with specific disease profiles along with education of enhanced public health prevention guidelines.

There are several limitations to this analysis. First, there is the agnostic nature of PheWAS, which can identify potentially spurious associations. While we feel that many of the top traits have been highlighted elsewhere and are biologically plausible, there is currently no process in place for rapidly discerning potentially novel from spurious associations[24] beyond extensive manual review and follow-up research, particularly for a novel disease. Second, many of the issues with utilizing EHR data for research purposes also applies here including inaccurate data from billing codes[20] and failure of physicians to report/record problems.[21] Third, the sample size for a PheWAS is still rather small to be able to identify statistically significant associations – particularly for mortality. Moreover, we did not distinguish between transfer patients (i.e., those who were diagnosed elsewhere and transferred to MM for treatment), who may have been sicker patients than the cohort diagnosed at MM. However, given that this is an emerging and novel disease, we feel it is important to identify suggestive associations so that future research and clinicians can potentially consider other conditions outside those that have been previously identified – namely, pulmonary and cardiovascular conditions – and to inspire follow-up studies in larger cohorts. For example, OpenSAFELY, a platform including primary care records of 17,728,392 adults in England (covering 40% of all patients),[27] shares many of our full-cohort conclusions (with consistent effect sizes), but has not published race-stratified PheWAS results. Finally, our analysis is scanning through each phenotype one at a time though they occur in a correlated and interactive manner. A richer multivariate model needs to be constructed with more complex features.

### 4.1. Conclusion

This work contributes to a new area of COVID-19 research that rigorously examines racial differences in disease prognosis with pre-existing conditions captured across the medical phenome. Moreover, we incorporated a census tract-level SES covariate, which are important to consider when comparing races [22]. We found several potentially novel diseases unexpectedly associated with different outcomes in the course of COVID-19 progression and that some disease profiles differ by race. For example, we provide additional evidence on the previously reported concern that patients with mental health disorders are at higher risk of infection and experience barriers in seeking treatment leading to poor prognosis [23]. We hope this exploratory effort will inspire hypothesis generation for future research that might result in targeted prevention and care as we are still combatting this pandemic. In this spirit, we have made all PheWAS results available for exploration here: https://cphds.sph.umich.edu/covidphewas/. We hope the summary data and the phenomic landscape for COVID-19 will help future replication and meta-analysis efforts.

## Data Availability

Data cannot be shared publicly due to patient confidentiality. The data underlying the results presented in the study are available from University of Michigan Data Office for Clinical & Translational Research for researchers who meet the criteria for access to confidential data.

## Abbreviations

Black: non-Hispanic Black/African American
EHR: electronic health record
ICD: International Classification of Disease
ICU: intensive care unit
MM: Michigan Medicine
NaNDA: National Neighborhood Data Archive
OR: odds ratio
PheWAS: phenome-wide association study
SES: socioeconomic status
White: non-Hispanic White

## Acknowledgments

Equal contribution of work

Maxwell Salvatore and Tian Gu contributed equally to this work

