## Supplementary Material for "A phenome-wide association study (PheWAS) of COVID-19 outcomes by race using the electronic health records data in Michigan Medicine"

*Control selection for susceptibility PheWAS.* Controls were used in the susceptibility models and not in the prognosis models. We created two sets of alive control samples from the MM patient database to compare and contrast the testing positive, one “unmatched control sample” consists of 29,383 randomly drawn patients and another 47,510 “matched control sample” using 1:3 frequency-matching on race, ethnicity, sex and age (above/below 50) based on a previous cohort of about 16,000 tested individuals. We used unmatched controls in the analysis with the full cohort (including all races) and matched controls in the race-stratified analysis. Matched controls were used in the race-stratified models for COVID-19 susceptibility as the proportion of non-Hispanic Whites (White) and non-Hispanic Black/African American (Black) in unmatched controls are not comparable to the stratified population under study.

There were 53,853 patients who were either tested for or diagnosed with COVID-19, 29,383 unmatched controls (n_unmatched_), and 47,510 matched controls (n_matched_) eligible for inclusion in this study. Of the 130,746 individuals eligible for inclusion, our study population comprised 123,344 individuals (n_tested_=47,862 [n_positive_=2,133]; n_unmatched_=28,505; n_matched_= 46,977) who had available International Classification of Disease (ICD; ninth and tenth editions) code data after applying the 14-day-prior to testing restriction to the EHR. Furthermore, a total of 1,813 qualified ICD-code-based phenotypes, referred to as PheWAS codes, were initially screened of which 1,363 had at least 10 occurrences in our COVID-19 positive cohort and were included in the analysis.

*Full cohort susceptibility.* For susceptibility, when comparing the positives and the unmatched controls, 929 traits were identified after applying Bonferroni correction, including 101 genitourinary, 97 circulatory system, 91 endocrine/metabolic, and 89 digestive PheWAS codes. This suggests that patients who tested positive were enriched for pre-existing conditions compared to the control population. As illustrated in **Figure S2A**, we found strong and positive associations with various comorbidities and COVID-19 positive diagnosis (e.g., shortness of breath [*P*=1.00x10^-127^], cough [*P*=1.00x10^-119^], pneumonia [*P*=1.00x10^-108^], nonspecific chest pain [*P*=7.94x10^-95^], vitamin D deficiency [*P*=1.26x10^-70^] and anxiety disorders [*P*=5.01x10^-53^]). Overall, the findings were consistent with previously identified COVID-19 risk factors (e.g., obesity [*P*=1.26x10^-95^] and diabetes mellitus [*P*=3.16x10^-64^] were associated with higher risk of being test positive) (**Table S3**).^19^ In contrast, the comparison between those who tested positive for COVID-19 and those who tested negative leads to counterintuitive findings (all 137 significant traits under Bonferroni correction showed protective effect, such as cataract [odds ratio [OR]=0.45, *P*=6.31x10^-7^] and asthma [OR=0.68, *P*=3.98x10^-18^]) contradicting findings in other COVID-19 studies.^20,21^ However, this can likely be explained by the applied eligibility criteria, e.g., a larger proportion of COVID-19-positive individuals became eligible due to symptom manifestation while COVID-19-negative individuals were tested due to Michigan’s priority testing criteria^22^ (without symptoms). This amplifies the need for choosing an appropriate control group.

*Race-stratified susceptibility analysis.* As shown in **Figure S2B**, we identified 871 traits in Whites, including 122 circulatory system, 120 digestive, 119 genitourinary, and 107 endocrine/metabolic diseases under Bonferroni correction. Examples of the top traits include cough (*P*=3.98x10^-84^), nonspecific chest pain (*P*=3.16x10^-68^), malaise and fatigue (*P*=2.00x10^-64^), abnormal glucose (*P*=1.58x10^-58^), and pain in joint (*P*=1.26x10^-56^). As illustrated in **Figure S2C**, we observed 584 significant traits in Blacks under Bonferroni correction, including 118 circulatory system, 110 genitourinary, 102 endocrine/metabolic, 99 digestive and 68 musculoskeletal diseases, where some of the top traits includes nonspecific chest pain (*P*=2.51x10^-37^), electrolyte imbalance (*P*=1.00x10^-36^), acute renal failure (*P*=1.26x10^-35^) and cardiac dysrhythmias (*P*=2.00x10^-33^).

**Figure S1**. Forest plots of traits associated with poorer prognosis in overall cohort. *A. Hospitalization. B.* *ICU admission/mortality. Odds ratios and 95% confidence intervals are shown for each trait whose PheWAS code is given in parentheses. Plots show child codes only. Parent codes found in Figure 3.*

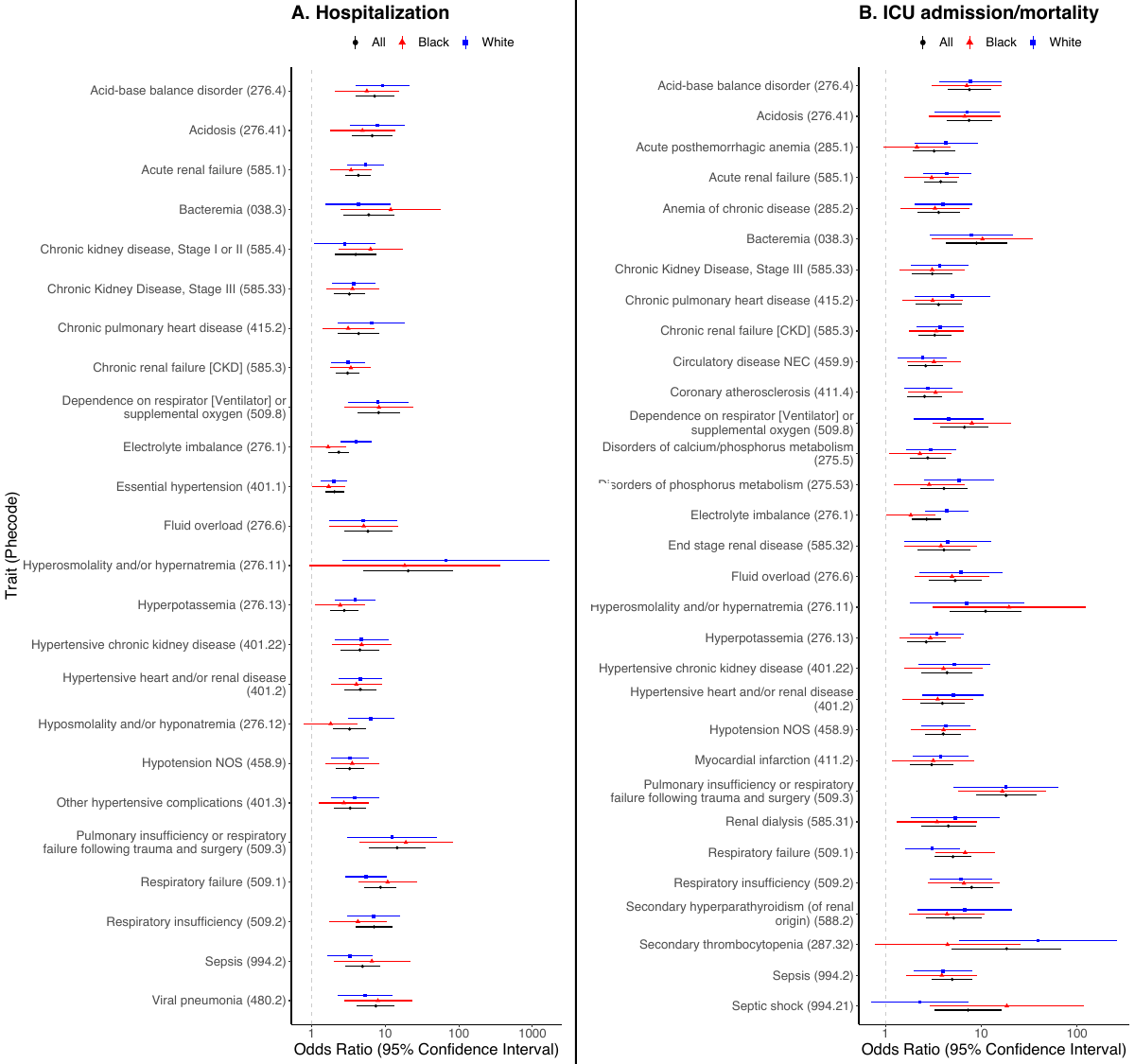

**Figure S2**. Manhattan plot showing the phenome-wide association between disease codes and testing positive for COVID-19. *Models are adjusted for age, sex, race (full cohort only), and four census tract-level socioeconomic indicators: proportion with less than high school education, proportion unemployed, proportion with annual income below the federal poverty level, and population density (persons per mile^2^). The x-axis are individual disease codes, color-coded by their corresponding disease category as described in the legend. The y-axis represents the -log_10_ transformed p-value of the association. The dashed, horizontal lines represent the p = 0.05 (in orange) and the Bonferroni corrected p-value (0.05 / number of tests; in red). Each point is represented by either an upward triangle indicating a positive association or a downward triangle indicating a negative association.*

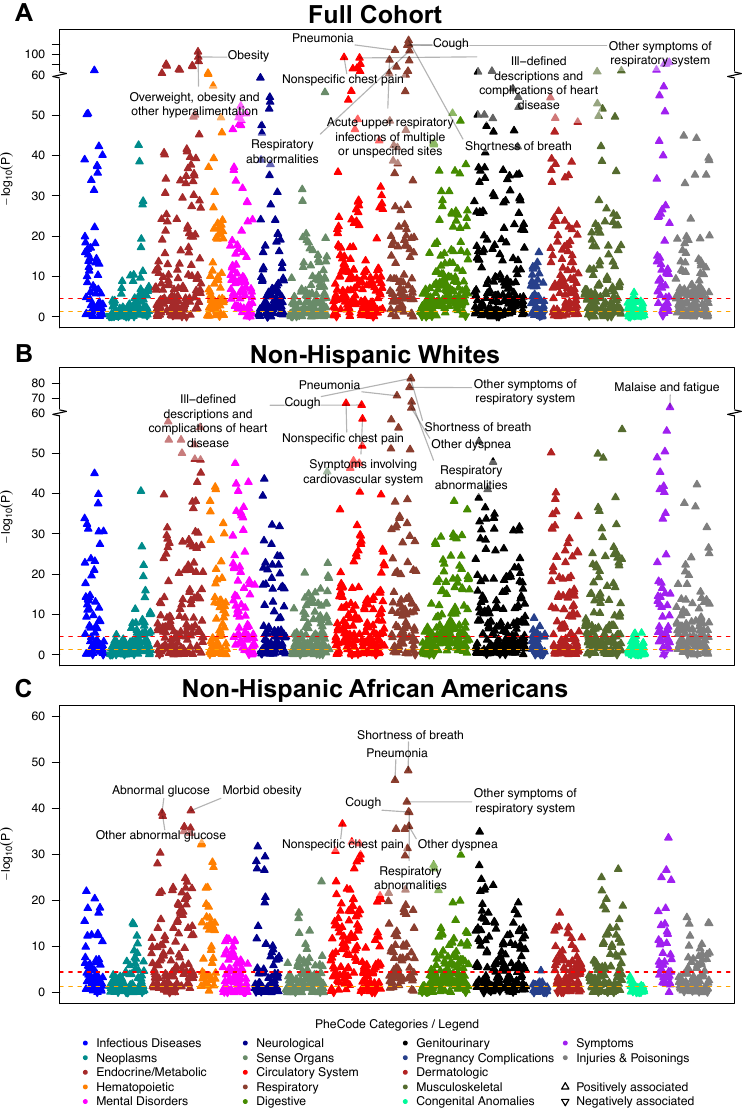

|  | **Definition** | **Sources** |
| --- | --- | --- |
| Age | Age of patient as of the data pull: 4/22/2020 | Electronic Health Record (EPIC) |
| Male | Gender of patient as reported. | Electronic Health Record (EPIC) |
| BMI | Excluded entries if (1) age at BMI measurement was missing or below 18 years, (2) height and/or weight were missing, (3) height measurements were below 69 cm or above 234 cm, (4) weight was above 400 kg, (5) BMI deviated more than one unit from BMI calculated from height and weight (BMI = weight [in kg] / height [in m]^2 ). Outliers for multiple values per person were defined as values that exceeded the median BMI +/- 3 x the median absolute deviation (MAD). Final BMI values was calculated as the median BMI of the remaining entries. | Derived from the Electronic Health Record |
| **Race/Ethnicity** |  | Patient Reported - Derived from the Electronic Health Record |
| White | If race was reported as "Caucasian" and ethnicity as "Hispanic or Latino" |  |
| Black | If race was reported as "African American" and ethnicity as "Hispanic or Latino" |  |
| Other / Known Ethnicity | If race was not reported as "African American" or "Caucasian"' and ethnicity was reported as "Non-Hispanic or Latino" or "Hispanic or Latino" |  |
| Other / Unknown Ethnicity | If race and/or ethnicity were missing |  |
| **SES** | Data defined by US census tract (based on residential address available in each patient’s EHR) for the year 2010 from the US Census and the American Community Survey (ACS). | The boundaries for the census tracts were normalized by 2010 tract boundaries using the Longitudinal Tract Data Base (Logan, Xu, and Stults, 2014). |
| Less than HS Education | Proportion of the Neighborhood Census Tract that has less than a High School Education |  |
| Unemployed | Proportion of the Neighborhood Census Tract age 16+ in the civilian labor force who are unemployed in 2010 |  |
| Annual Income Below FPL | Proportion of the Neighborhood Census Tract with annual income below the federal poverty level in 2010 |  |
| Persons Per Mile^2^ | Population Density of the neighborhood that the patient lives in. |  |
| **COVID-19 Outcomes** |  | Derived from RDW's COVID Registry |
| COVID-19 Tested | Patients who were tested for COVID-19 at the time of data pull. |  |
| COVID-19 Positive | Patients who tested positive at least once for COVID-19 |  |
| COVID-19 Negative | Patients who always tested negative for COVID-19 |  |
| Not-Hospitalized | Patients in the positive COVID cohort who have no inpatient stays after 3/5/2020 |  |
| Hospitalized | Patients in the positive COVID cohort who checked in as an inpatient after 3/5/2020 at least once. |  |
| ICU | Patients in the positive COVID cohort who checked into the ICU during their inpatient stay after 3/5/2020. |  |
| Deceased | Patients in the cohort who have died based on the Electronic Health Record. | Electronic Health Record (EPIC) |
| **Control Groups** |  |  |
| Unmatched Control | Randomly picked cohort of patients who are not part of the COVID-19 cohort (Tested, Positive, Negative), who are alive and who have had an encounter in MM (Inpatient, Outpatient or Emergency) since 2012-04-23 | Derived from the Electronic Health Record |
| Matched Control | Randomly picked cohort of patients matched by Age (<=50, >50), Race/Ethnicity, Gender to be 3 times the COVID cohort for each group. Find attached table for frequency matching. |  |

| **Full Cohort** | | Main Analysis | | | | | | | Sensitivity Analysis | | | | | | |
| --- | --- | --- | --- | --- | --- | --- | --- | --- | --- | --- | --- | --- | --- | --- | --- |
|  | | Tested (1)  vs. Controls (0) | Positive (1) vs.  Controls (0) | Positive (1) vs. Negative (0) | Hospitalized (1)  vs. Not (0) | | ICU (1) vs. Not (0) | Deceased (1) vs. Alive (0) | Positive (1) vs.  Controls (0) | | Positive (1) vs. Negative (0) | Hospitalized (1)  vs. Not (0) | | ICU (1) vs. Not (0) | Deceased (1) vs. Alive (0) |
| Not hospitalized | | 1 | 1 | 1 | 0 | | 0 | 0 | 1 | | 1 | 0 | | 0 | 0 |
| Not hospitalized / Deceased | | 1 | 1 | 1 | 1 | | 1 | 1 | 1 | | 1 | 1 | | 1 | 1 |
| Still hospitalized | | 1 | 1 | 1 | 1 | | 0 | 0 | NA | | NA | NA | | NA | NA |
| Hospitalized / Discharged | | 1 | 1 | 1 | 1 | | 0 | 0 | 1 | | 1 | 1 | | 0 | 0 |
| Hospitalized / Deceased | | 1 | 1 | 1 | 1 | | 1 | 1 | 1 | | 1 | 1 | | 1 | 1 |
| Still in ICU | | 1 | 1 | 1 | 1 | | 1 | 0 | NA | | NA | NA | | NA | NA |
| ICU / Discharged | | 1 | 1 | 1 | 1 | | 1 | 0 | 1 | | 1 | 1 | | 1 | 0 |
| ICU / Deceased | | 1 | 1 | 1 | 1 | | 1 | 1 | 1 | | 1 | 1 | | 1 | 1 |
| Matched Controls | | NA | NA | NA | NA | | NA | NA | NA | | NA | NA | | NA | NA |
| Unmatched Controls | | 0 | 0 | NA | NA | | NA | NA | 0 | | NA | NA | | NA | NA |
| Tested Negative | | 1 | NA | 0 | NA | | NA | NA | NA | | 0 | NA | | NA | NA |
| Adjustment 1 | | Age, Gender, Race/Ethnicity, Persons Per Mile^2^ | | | Age, Gender, Race/Ethnicity | | | | Age, Gender, Race/Ethnicity, Persons Per Mile^2^ | | | Age, Gender, Race/Ethnicity | | | |
| Adjustment 2 | | Adjustment 1 + <HS education, Unemployed, Annual Income Below FPL | | | | | | | | | | | | | |
| Adjustment 3 | | Adjustment 2 + Comorbidity Score | | | | | | | | | | | | | |
| **Race/ethnicity Stratified** | Positive (1) vs. Controls (0) | | | | | Hospitalized (1) vs. Not (0) | | | | ICU (1) vs. Not (0) | | | Dead (1) vs Alive (0) | | |
| Not hospitalized | 1 | | | | | 0 | | | | 0 | | | 0 | | |
| Not hospitalized / Deceased | 1 | | | | | 1 | | | | 1 | | | 1 | | |
| Still hospitalized | 1 | | | | | 1 | | | | 0 | | | 0 | | |
| Hospitalized / Discharged | 1 | | | | | 1 | | | | 0 | | | 0 | | |
| Hospitalized / Deceased | 1 | | | | | 1 | | | | 1 | | | 1 | | |
| Still in ICU | 1 | | | | | 1 | | | | 1 | | | 0 | | |
| ICU / Discharged | 1 | | | | | 1 | | | | 1 | | | 0 | | |
| ICU / Deceased | 1 | | | | | 1 | | | | 1 | | | 1 | | |
| Matched Controls | 0 | | | | | NA | | | | NA | | | NA | | |
| Unmatched Controls | NA | | | | | NA | | | | NA | | | NA | | |
| Tested Negative | NA | | | | | NA | | | | NA | | | NA | | |
| Adjustment 1 | Persons Per Mile^2^ | | | | | Age, Gender | | | | | | | | | |
| Adjustment 2 | Adjustment 1 + <HS education, Unemployed, Annual Income Below FPL | | | | | | | | | | | | | | |
| Adjustment 3 | Adjustment 2 + Comorbidity Score | | | | | | | | | | | | | | |

| **Non-Hispanic Whites** | | | | | | | | |
| --- | --- | --- | --- | --- | --- | --- | --- | --- |
| **Variable** | **Statistics** | **Tested** | **Tested Negative** | **Tested Positive** | **Hospitalized** | **ICU** | **Deceased** | **Unmatched Controls** |
| n |  | 38977 | 37566 | 1411 | 326 | 172 | 56 | 18529 |
| Age | mean (SD)  median (IQR) | 46.1 (23.3)  49 (37) | 46 (23.4)  49 (37) | 47 (20.3)  49 (34) | 59.9 (18.1)  63 (22) | 59.4 (18.5)  63 (22) | 72.2 (15.7)  77 (18.8) | 43 (24.5)  43 (42) |
| AgeCategory | n (%) |  |  |  |  |  |  |  |
| [0,18) |  | 5020 (12.9) | 4951 (13.2) | 69 (4.9) | 7 (2.1) | 5 (2.9) | 0 (0) | 3450 (18.6) |
| [18,35) |  | 8304 (21.3) | 7929 (21.1) | 375 (26.6) | 31 (9.5) | 17 (9.9) | 3 (5.4) | 4232 (22.8) |
| [35,50) |  | 6345 (16.3) | 6058 (16.1) | 287 (20.3) | 40 (12.3) | 20 (11.6) | 3 (5.4) | 2816 (15.2) |
| [50,65) |  | 9063 (23.3) | 8702 (23.2) | 361 (25.6) | 100 (30.7) | 51 (29.7) | 8 (14.3) | 3560 (19.2) |
| [65,80) |  | 8177 (21) | 7928 (21.1) | 249 (17.6) | 105 (32.2) | 61 (35.5) | 18 (32.1) | 3320 (17.9) |
| [80,100) |  | 2068 (5.3) | 1998 (5.3) | 70 (5) | 43 (13.2) | 18 (10.5) | 24 (42.9) | 1151 (6.2) |
| Gender | (Male; %) | 17327 (44.5) | 16672 (44.4) | 655 (46.4) | 196 (60.1) | 114 (66.3) | 38 (67.9) | 8606 (46.5) |
| Primary Care in MM | (Yes; %) | 23813 (61.1) | 22935 (61.1) | 878 (62.2) | 139 (42.6) | 81 (47.1) | 22 (39.3) | 2939 (15.9) |
| BMI | mean (SD) | 29 (7.3) | 29 (7.3) | 29.6 (7.4) | 31.2 (8.2) | 30.5 (8.4) | 30 (6.9) | 28.5 (7.3) |
| BMIcategory | n (%) |  |  |  |  |  |  |  |
| [0,18.5) |  | 571 (1.8) | 559 (1.8) | 12 (1) | 4 (1.3) | 4 (2.4) | 1 (1.9) | 243 (2) |
| [18.5,25) |  | 9718 (29.8) | 9366 (29.9) | 352 (28.4) | 57 (18.3) | 37 (22.4) | 8 (14.8) | 3826 (32) |
| [25,30) |  | 10187 (31.3) | 9799 (31.3) | 388 (31.3) | 103 (33) | 56 (33.9) | 23 (42.6) | 3781 (31.7) |
| [30,200) |  | 12085 (37.1) | 11598 (37) | 487 (39.3) | 148 (47.4) | 68 (41.2) | 22 (40.7) | 4090 (34.3) |
| Smoker | (Ever; %) | 14197 (38.3) | 13773 (38.5) | 424 (33.2) | 134 (45.4) | 73 (48.3) | 22 (55) | 5378 (33.3) |
| SmokingStatus | n (%) |  |  |  |  |  |  |  |
| Never |  | 22832 (61.7) | 21978 (61.5) | 854 (66.8) | 161 (54.6) | 78 (51.7) | 18 (45) | 10763 (66.7) |
| Past |  | 10999 (29.7) | 10630 (29.7) | 369 (28.9) | 120 (40.7) | 66 (43.7) | 21 (52.5) | 3707 (23) |
| Current |  | 3198 (8.6) | 3143 (8.8) | 55 (4.3) | 14 (4.7) | 7 (4.6) | 1 (2.5) | 1671 (10.4) |
| Drinker | (Yes; %) | 20623 (70.9) | 19894 (70.9) | 729 (69.7) | 151 (69.3) | 78 (66.7) | 22 (71) | 6718 (57.3) |
| NDI | mean (SD) | 0.09 (0.06) | 0.09 (0.06) | 0.09 (0.06) | 0.09 (0.06) | 0.1 (0.06) | 0.1 (0.05) | 0.1 (0.06) |
| PersonsPerSquareMile | mean (SD) | 2106.4 (2263.9) | 2094.9 (2263.5) | 2405.7 (2252.8) | 2711.6 (2228.1) | 2895.7 (2343.9) | 2880.7 (1950.8) | 2057.5 (2316.2) |
| ComorbidityScore | mean (SD) | 2.3 (1.5) | 2.3 (1.5) | 2.3 (1.5) | 3.1 (1.6) | 3.3 (1.7) | 3.9 (1.6) | 1.4 (1.2) |

| **Non-Hispanic African Americans** | | | | | | | | |
| --- | --- | --- | --- | --- | --- | --- | --- | --- |
| **Variable** | **Statistics** | **Tested** | **Tested Negative** | **Tested Positive** | **Hospitalized** | **ICU** | **Deceased** | **Unmatched Controls** |
| n |  | 5763 | 5117 | 646 | 265 | 139 | 42 | 2165 |
| Age | mean (SD)  median (IQR) | 42.8 (21.7)  44 (34) | 42.1 (22)  43 (35) | 48.6 (18.6)  49 (27) | 56.8 (17.1)  58 (24) | 57.8 (17)  59 (22.5) | 67.2 (11.4)  66.5 (17.5) | 36.8 (23.2)  34 (38) |
| AgeCategory | n (%) |  |  |  |  |  |  |  |
| [0,18) |  | 697 (12.1) | 675 (13.2) | 22 (3.4) | 5 (1.9) | 4 (2.9) | 0 (0) | 547 (25.3) |
| [18,35) |  | 1487 (25.8) | 1355 (26.5) | 132 (20.4) | 21 (7.9) | 11 (7.9) | 0 (0) | 541 (25) |
| [35,50) |  | 1173 (20.4) | 1001 (19.6) | 172 (26.6) | 63 (23.8) | 23 (16.5) | 3 (7.1) | 376 (17.4) |
| [50,65) |  | 1340 (23.3) | 1157 (22.6) | 183 (28.3) | 83 (31.3) | 50 (36) | 15 (35.7) | 384 (17.7) |
| [65,80) |  | 890 (15.4) | 784 (15.3) | 106 (16.4) | 67 (25.3) | 39 (28.1) | 15 (35.7) | 249 (11.5) |
| [80,100) |  | 176 (3.1) | 145 (2.8) | 31 (4.8) | 26 (9.8) | 12 (8.6) | 9 (21.4) | 68 (3.1) |
| Gender | (Male; %) | 2336 (40.5) | 2067 (40.4) | 269 (41.6) | 140 (52.8) | 81 (58.3) | 25 (59.5) | 1011 (46.7) |
| Primary Care in MM | (Yes; %) | 3284 (57) | 2973 (58.1) | 311 (48.1) | 83 (31.3) | 34 (24.5) | 9 (21.4) | 431 (19.9) |
| BMI | mean (SD) | 31.5 (8.8) | 31.1 (8.6) | 34 (9.9) | 34.6 (12.1) | 35.4 (14.3) | 33.1 (6.5) | 30.7 (8.3) |
| BMIcategory | n (%) |  |  |  |  |  |  |  |
| [0,18.5) |  | 100 (2.1) | 92 (2.2) | 8 (1.4) | 5 (2) | 0 (0) | 0 (0) | 22 (1.7) |
| [18.5,25) |  | 996 (20.6) | 940 (22.1) | 56 (9.6) | 20 (7.8) | 13 (9.8) | 3 (7.1) | 298 (23.7) |
| [25,30) |  | 1327 (27.4) | 1183 (27.8) | 144 (24.8) | 70 (27.5) | 36 (27.1) | 13 (31) | 357 (28.3) |
| [30,200) |  | 2421 (50) | 2048 (48) | 373 (64.2) | 160 (62.7) | 84 (63.2) | 26 (61.9) | 583 (46.3) |
| Smoker | (Ever; %) | 2011 (37.4) | 1831 (38) | 180 (32.2) | 83 (36.7) | 43 (41) | 19 (70.4) | 517 (28.3) |
| SmokingStatus | n (%) |  |  |  |  |  |  |  |
| Never |  | 3367 (62.6) | 2988 (62) | 379 (67.8) | 143 (63.3) | 62 (59) | 8 (29.6) | 1312 (71.7) |
| Past |  | 1350 (25.1) | 1200 (24.9) | 150 (26.8) | 77 (34.1) | 41 (39) | 19 (70.4) | 304 (16.6) |
| Current |  | 661 (12.3) | 631 (13.1) | 30 (5.4) | 6 (2.7) | 2 (1.9) | 0 (0) | 213 (11.6) |
| Drinker | (Yes; %) | 2393 (58.4) | 2134 (58) | 259 (62.3) | 81 (57) | 37 (57.8) | 12 (60) | 575 (44.7) |
| NDI | mean (SD) | 0.18 (0.1) | 0.18 (0.1) | 0.19 (0.11) | 0.2 (0.11) | 0.21 (0.11) | 0.21 (0.1) | 0.19 (0.11) |
| PersonsPerSquareMile | mean (SD) | 3594.3 (2406.4) | 3547.9 (2407.2) | 3937.7 (2374.8) | 4439.2 (2482) | 4555.3 (2472.3) | 5050.3 (2260.1) | 3541.3 (2604.8) |
| ComorbidityScore | mean (SD) | 2.5 (1.6) | 2.5 (1.6) | 2.7 (1.6) | 3.2 (1.6) | 3.4 (1.6) | 4.3 (1.4) | 1.4 (1.3) |

| Trait (Phecode) | Category | Rank  All/NHW/NHB | ALL  OR (95% CI) P-value | NHW  OR (95% CI) P-value | NHB  OR (95% CI) P-value |
| --- | --- | --- | --- | --- | --- |
| Viral infection (079) | infectious diseases | 27/41/79 | 3.91 (3.36, 4.55) 1.36e-69 | 3.88 (3.22, 4.68) 9.2e-46 | 3.35 (2.61, 4.31) 3.72e-21 |
| Diabetes mellitus (250) | endocrine/metabolic | 35/59/35 | 3.34 (2.91, 3.84) 3.32e-64 | 3.19 (2.69, 3.79) 2.22e-40 | 3.39 (2.73, 4.2) 9e-29 |
| Type 2 diabetes with renal manifestations (250.22) | endocrine/metabolic | 108/129/26 | 6.96 (5.24, 9.24) 5.65e-41 | 7.96 (5.58, 11.4) 2.71e-30 | 8.46 (5.89, 12.1) 4.55e-31 |
| Abnormal glucose (250.4) | endocrine/metabolic | 17/12/7 | 4.73 (4.03, 5.54) 1.42e-81 | 5.04 (4.14, 6.13) 1.59e-58 | 5.5 (4.27, 7.09) 9.45e-40 |
| Other abnormal glucose (250.42) | endocrine/metabolic | 19/18/8 | 4.9 (4.15, 5.78) 2.96e-78 | 5.11 (4.16, 6.28) 5.17e-54 | 5.71 (4.39, 7.41) 4.69e-39 |
| Vitamin deficiency (261) | endocrine/metabolic | 24/17/65 | 4.15 (3.55, 4.85) 1.34e-71 | 4.69 (3.85, 5.7) 5.05e-54 | 3.57 (2.77, 4.59) 5.63e-23 |
| Vitamin D deficiency (261.4) | endocrine/metabolic | 25/26/55 | 4.23 (3.61, 4.96) 1.28e-70 | 4.65 (3.8, 5.69) 9.15e-51 | 3.68 (2.85, 4.74) 1.46e-23 |
| Disorders of fluid, electrolyte, and acid-base balance (276) | endocrine/metabolic | 18/20/15 | 4.17 (3.59, 4.84) 1.04e-78 | 4.4 (3.64, 5.31) 8.24e-53 | 4.54 (3.58, 5.75) 9.8e-36 |
| Electrolyte imbalance (276.1) | endocrine/metabolic | 20/30/11 | 5.25 (4.41, 6.25) 2.76e-77 | 5.21 (4.18, 6.48) 3.21e-49 | 5.78 (4.4, 7.58) 9.91e-37 |
| Hyperpotassemia (276.13) | endocrine/metabolic | 67/66/48 | 6.54 (5.11, 8.37) 2.33e-50 | 6.98 (5.21, 9.34) 7.15e-39 | 7.41 (5.09, 10.8) 1.62e-25 |
| Hypovolemia (276.5) | endocrine/metabolic | 63/43/53 | 4.46 (3.66, 5.42) 7.74e-51 | 5.35 (4.22, 6.78) 1.62e-43 | 4.94 (3.64, 6.71) 1.07e-24 |
| Overweight, obesity and other hyperalimentation (278) | endocrine/metabolic | 6/13/17 | 4.15 (3.65, 4.71) 3.18e-106 | 3.81 (3.23, 4.49) 3.8e-57 | 3.89 (3.14, 4.83) 2.77e-35 |
| Obesity (278.1) | endocrine/metabolic | 7/31/12 | 4.19 (3.66, 4.79) 1.21e-95 | 3.75 (3.14, 4.47) 4.71e-49 | 4.11 (3.3, 5.12) 1.89e-36 |
| Morbid obesity (278.11) | endocrine/metabolic | 12/40/4 | 5.56 (4.69, 6.59) 4.09e-87 | 4.91 (3.94, 6.11) 7.86e-46 | 5.54 (4.3, 7.13) 3.23e-40 |
| Iron deficiency anemias (280) | hematopoietic | 34/65/22 | 4.94 (4.1, 5.94) 2.38e-64 | 4.9 (3.86, 6.23) 7.1e-39 | 5.11 (3.91, 6.68) 7.35e-33 |
| Iron deficiency anemias, unspecified or not due to blood loss (280.1) | hematopoietic | 37/81/20 | 4.93 (4.08, 5.94) 1.52e-62 | 4.81 (3.76, 6.14) 3.1e-36 | 5.2 (3.97, 6.8) 3.56e-33 |
| Other anemias (285) | hematopoietic | 40/48/58 | 3.2 (2.78, 3.69) 4.8e-58 | 3.54 (2.95, 4.24) 2.38e-42 | 3.13 (2.5, 3.91) 1.89e-23 |
| Anemia of chronic disease (285.2) | hematopoietic | 114/49/134 | 6.66 (5.03, 8.82) 5.44e-40 | 9.2 (6.67, 12.7) 9.05e-42 | 5.75 (3.83, 8.65) 4.29e-17 |
| Diseases of white blood cells (288) | hematopoietic | 69/99/34 | 4.5 (3.69, 5.48) 3.97e-50 | 4.58 (3.57, 5.87) 2.46e-33 | 5.73 (4.22, 7.78) 4.75e-29 |
| Elevated white blood cell count (288.2) | hematopoietic | 88/125/38 | 4.73 (3.82, 5.85) 2.2e-46 | 4.85 (3.71, 6.35) 1.11e-30 | 6.2 (4.47, 8.6) 6.44e-28 |
| Other specified nonpsychotic and/or transient mental disorders (291) | mental disorders | 83/34/256 | 4.9 (3.95, 6.08) 3.45e-47 | 6.39 (4.98, 8.2) 2.91e-48 | 3.55 (2.47, 5.11) 8.79e-12 |
| Alteration of consciousness (291.8) | mental disorders | 136/45/321 | 5.05 (3.94, 6.48) 4.2e-37 | 7.26 (5.48, 9.63) 4.14e-43 | 3.68 (2.43, 5.59) 9.01e-10 |
| Adjustment reaction (304) | mental disorders | 59/44/406 | 5.09 (4.11, 6.29) 4.23e-51 | 5.89 (4.58, 7.57) 1.76e-43 | 2.68 (1.87, 3.86) 9.56e-08 |
| Sleep disorders (327) | neurological | 39/60/32 | 3.07 (2.68, 3.51) 5.36e-60 | 3.15 (2.66, 3.73) 3.82e-40 | 3.59 (2.87, 4.49) 2.84e-29 |
| Sleep apnea (327.3) | neurological | 117/233/40 | 2.91 (2.48, 3.41) 1.14e-39 | 2.77 (2.25, 3.4) 2.85e-22 | 3.96 (3.09, 5.07) 1.14e-27 |
| Obstructive sleep apnea (327.32) | neurological | 80/144/23 | 3.59 (3.02, 4.26) 3.45e-48 | 3.48 (2.8, 4.34) 5.57e-29 | 5.05 (3.86, 6.6) 1.99e-32 |
| Insomnia (327.4) | neurological | 89/42/168 | 3.94 (3.26, 4.75) 2.63e-46 | 4.74 (3.81, 5.89) 2.51e-44 | 3.72 (2.68, 5.17) 4.32e-15 |
| Pain (338) | neurological | 56/154/44 | 2.78 (2.44, 3.17) 2.69e-52 | 2.59 (2.19, 3.07) 3.66e-28 | 3.34 (2.68, 4.15) 2.51e-27 |
| Chronic pain (338.2) | neurological | 47/102/30 | 3.31 (2.85, 3.84) 3.17e-55 | 3.22 (2.66, 3.9) 6.19e-33 | 3.99 (3.15, 5.06) 3.32e-30 |
| Dizziness and giddiness (Light-headedness and vertigo) (386.9) | sense organs | 45/38/52 | 3.6 (3.07, 4.22) 2.19e-56 | 4.1 (3.38, 4.98) 3.64e-46 | 3.89 (3.01, 5.05) 8.19e-25 |
| Hypertensive heart and/or renal disease (401.2) | circulatory system | 142/77/25 | 4.97 (3.87, 6.37) 1.39e-36 | 6.44 (4.83, 8.59) 9.61e-37 | 6.94 (5.01, 9.61) 2.16e-31 |
| Nonspecific chest pain (418) | circulatory system | 8/6/9 | 4 (3.5, 4.56) 7.9e-95 | 4.36 (3.69, 5.16) 3.13e-67 | 4.13 (3.32, 5.13) 2.25e-37 |
| Cardiac conduction disorders (426) | circulatory system | 50/37/61 | 3.56 (3.04, 4.18) 1.53e-54 | 4.13 (3.41, 5.01) 5.47e-47 | 3.53 (2.75, 4.53) 3.53e-23 |
| Abnormal electrocardiogram [ECG] [EKG] (426.7) | circulatory system | 43/32/63 | 3.82 (3.24, 4.51) 8.91e-57 | 4.49 (3.67, 5.49) 7.25e-49 | 3.65 (2.82, 4.72) 5.11e-23 |
| Cardiac dysrhythmias (427) | circulatory system | 23/36/19 | 3.46 (3.02, 3.96) 7.36e-73 | 3.52 (2.97, 4.18) 6.97e-48 | 3.94 (3.15, 4.93) 1.89e-33 |
| Other specified cardiac dysrhythmias (427.3) | circulatory system | 82/103/41 | 3.71 (3.11, 4.44) 3.4e-47 | 3.97 (3.17, 4.98) 7.17e-33 | 4.63 (3.51, 6.1) 1.2e-27 |
| Tachycardia NOS (427.7) | circulatory system | 22/35/33 | 6.01 (4.96, 7.29) 4.02e-74 | 5.96 (4.69, 7.57) 3.15e-48 | 5.57 (4.12, 7.52) 4.26e-29 |
| Ill-defined descriptions and complications of heart disease (429) | circulatory system | 9/7/21 | 4.26 (3.71, 4.89) 8.12e-94 | 4.51 (3.8, 5.36) 4.92e-66 | 3.99 (3.18, 5.01) 4.48e-33 |
| Abnormal function study of cardiovascular system (429.2) | circulatory system | 31/21/31 | 5.04 (4.2, 6.05) 6.76e-68 | 5.62 (4.5, 7.02) 1.71e-52 | 4.86 (3.69, 6.39) 1.25e-29 |
| Symptoms involving cardiovascular system (429.3) | circulatory system | 16/10/27 | 4.43 (3.8, 5.15) 6e-82 | 4.69 (3.89, 5.65) 3.15e-59 | 4.37 (3.4, 5.61) 1.21e-30 |
| Acute sinusitis (464) | respiratory | 36/22/86 | 5.08 (4.21, 6.14) 1.69e-63 | 5.51 (4.42, 6.88) 7.81e-52 | 5.06 (3.59, 7.12) 1.73e-20 |
| Acute upper respiratory infections of multiple or unspecified sites (465) | respiratory | 10/11/72 | 4.04 (3.52, 4.62) 7.44e-90 | 4 (3.38, 4.73) 4.65e-59 | 3.04 (2.43, 3.81) 2.57e-22 |
| Pneumonia (480) | respiratory | 4/4/2 | 5.82 (4.98, 6.8) 2.47e-108 | 6.08 (4.99, 7.4) 2.77e-72 | 6.8 (5.23, 8.83) 8.86e-47 |
| Viral pneumonia (480.2) | respiratory | 21/14/13 | 85.7 (53.5, 137) 1.87e-76 | 51.1 (31.4, 82.9) 5.43e-57 | 60.5 (31.9, 115) 2.94e-36 |
| Respiratory failure, insufficiency, arrest (509) | respiratory | 44/98/14 | 4.86 (3.99, 5.9) 1.3e-56 | 4.95 (3.81, 6.42) 2.25e-33 | 6.67 (4.96, 8.97) 3.2e-36 |
| Respiratory failure (509.1) | respiratory | 38/64/29 | 7.49 (5.89, 9.52) 6.52e-61 | 7.75 (5.7, 10.5) 2.99e-39 | 7.97 (5.59, 11.4) 2.21e-30 |
| Other symptoms of respiratory system (512) | respiratory | 3/3/3 | 4.4 (3.88, 4.99) 3.95e-119 | 4.75 (4.04, 5.59) 5.48e-78 | 4.56 (3.66, 5.68) 4.23e-42 |
| Painful respiration (512.2) | respiratory | 30/23/24 | 8.82 (6.91, 11.3) 5.39e-68 | 9.91 (7.36, 13.3) 1.17e-51 | 9.03 (6.26, 13) 4.83e-32 |
| Shortness of breath (512.7) | respiratory | 1/2/1 | 6.25 (5.38, 7.25) 1.89e-127 | 6.72 (5.55, 8.15) 6.83e-84 | 6.28 (4.91, 8.02) 6.6e-49 |
| Cough (512.8) | respiratory | 2/1/5 | 5.37 (4.66, 6.19) 2.39e-119 | 6 (5.01, 7.19) 3.73e-84 | 5.35 (4.17, 6.86) 6.44e-40 |
| Other dyspnea (512.9) | respiratory | 11/9/10 | 4.74 (4.06, 5.52) 1.95e-88 | 5.43 (4.46, 6.6) 4.86e-64 | 5.32 (4.11, 6.89) 8.9e-37 |
| Respiratory abnormalities (513) | respiratory | 5/5/6 | 4.38 (3.84, 4.99) 5.64e-108 | 4.47 (3.78, 5.29) 2.88e-68 | 4.39 (3.53, 5.47) 7.42e-40 |
| Diseases of esophagus (530) | digestive | 96/148/36 | 2.61 (2.28, 2.99) 7.46e-44 | 2.63 (2.21, 3.12) 1.37e-28 | 3.55 (2.84, 4.45) 2.07e-28 |
| Esophagitis, GERD and related diseases (530.1) | digestive | 97/155/39 | 2.62 (2.29, 3.01) 1.89e-43 | 2.63 (2.22, 3.13) 3.7e-28 | 3.53 (2.82, 4.43) 9.69e-28 |
| GERD (530.11) | digestive | 95/156/42 | 2.66 (2.31, 3.05) 7.28e-44 | 2.66 (2.23, 3.16) 3.96e-28 | 3.54 (2.82, 4.45) 1.26e-27 |
| Abnormal serum enzyme levels (573.9) | digestive | 76/128/28 | 4.24 (3.5, 5.14) 2.76e-49 | 4.26 (3.32, 5.46) 2.28e-30 | 5.56 (4.15, 7.45) 1.31e-30 |
| Renal failure (585) | genitourinary | 64/62/37 | 3.75 (3.15, 4.46) 8.46e-51 | 4.19 (3.39, 5.19) 1.25e-39 | 4.05 (3.16, 5.2) 2.8e-28 |
| Acute renal failure (585.1) | genitourinary | 33/19/16 | 6.2 (5.04, 7.63) 7.5e-67 | 7.28 (5.65, 9.37) 1.38e-53 | 6.6 (4.9, 8.88) 1.21e-35 |
| Chronic Kidney Disease, Stage III (585.33) | genitourinary | 166/138/49 | 4.8 (3.73, 6.18) 3.72e-34 | 5.6 (4.15, 7.55) 1.66e-29 | 6.64 (4.65, 9.49) 2.26e-25 |
| Hematuria (593) | genitourinary | 81/50/123 | 4.3 (3.54, 5.24) 4.81e-48 | 5.12 (4.04, 6.48) 9.51e-42 | 4.33 (3.11, 6.04) 4.61e-18 |
| Other symptoms/disorders or the urinary system (599) | genitourinary | 29/33/54 | 3.33 (2.91, 3.82) 4.09e-68 | 3.57 (3.01, 4.23) 1.75e-48 | 3.26 (2.59, 4.1) 4.84e-24 |
| Noninflammatory female genital disorders (619) | genitourinary | 42/119/155 | 4.35 (3.63, 5.21) 2.5e-57 | 4.05 (3.2, 5.11) 1.51e-31 | 3.22 (2.42, 4.27) 7.45e-16 |
| Disorders of menstruation and other abnormal bleeding from female genital tract (626) | genitourinary | 48/120/197 | 4.3 (3.58, 5.16) 3.2e-55 | 4.11 (3.24, 5.22) 1.67e-31 | 3.08 (2.3, 4.13) 5.12e-14 |
| Superficial cellulitis and abscess (681) | dermatologic | 49/25/195 | 3.97 (3.34, 4.73) 4.12e-55 | 4.76 (3.88, 5.83) 7.23e-51 | 3.13 (2.33, 4.22) 4.43e-14 |
| Peripheral enthesopathies and allied syndromes (726) | musculoskeletal | 32/27/47 | 4.09 (3.49, 4.8) 3.87e-67 | 4.44 (3.65, 5.4) 1.63e-50 | 4.22 (3.23, 5.53) 1.08e-25 |
| Other disorders of soft tissues (729) | musculoskeletal | 55/29/81 | 5.71 (4.56, 7.14) 2.27e-52 | 7.52 (5.76, 9.83) 1.6e-49 | 5.48 (3.84, 7.81) 5.13e-21 |
| Symptoms and disorders of the joints (741) | musculoskeletal | 68/61/43 | 3.39 (2.89, 3.98) 2.72e-50 | 3.85 (3.15, 4.7) 1.1e-39 | 4.2 (3.24, 5.44) 1.42e-27 |
| Pain in joint (745) | musculoskeletal | 28/15/100 | 3.05 (2.69, 3.46) 6.29e-69 | 3.71 (3.15, 4.36) 1.41e-56 | 2.69 (2.17, 3.33) 1.54e-19 |
| Back pain (760) | symptoms | 26/28/119 | 3.21 (2.83, 3.66) 1.82e-70 | 3.42 (2.91, 4.03) 1.39e-49 | 2.63 (2.12, 3.27) 2.79e-18 |
| Swelling of limb (771.1) | symptoms | 46/39/46 | 4.52 (3.74, 5.45) 1.34e-55 | 5.31 (4.22, 6.68) 4.99e-46 | 4.55 (3.43, 6.04) 1.04e-25 |
| Edema (782.3) | symptoms | 110/47/161 | 3.75 (3.09, 4.55) 9.52e-41 | 4.75 (3.8, 5.94) 1.74e-42 | 3.47 (2.56, 4.7) 1.24e-15 |

| Trait (Phecode) | Category | Rank  All/NHW/NHB | ALL  OR (95% CI) P-value | NHW  OR (95% CI) P-value | NHB  OR (95% CI) P-value |
| --- | --- | --- | --- | --- | --- |
| Septicemia (038) | infectious diseases | 12/62/8 | 4.32 (2.65, 7.04) 4.08e-09 | 2.95 (1.55, 5.61) 0.000948 | 7.25 (2.6, 20.2) 0.000151 |
| Bacteremia (038.3) | infectious diseases | 40/117/18 | 5.98 (2.69, 13.3) 1.17e-05 | 4.31 (1.56, 11.9) 0.00471 | 11.9 (2.48, 57.4) 0.002 |
| Streptococcus infection (041.2) | infectious diseases | 46/43/51 | 4.69 (2.24, 9.84) 4.26e-05 | 6.66 (2.36, 18.8) 0.000346 | 4.11 (1.24, 13.7) 0.0211 |
| Protein-calorie malnutrition (260) | endocrine/metabolic | 79/31/518 | 2.54 (1.49, 4.33) 0.000584 | 4.02 (1.96, 8.21) 0.000138 | 1.21 (0.494, 2.98) 0.672 |
| Disorders of carbohydrate transport and metabolism (271) | endocrine/metabolic | 155/248/36 | 0.537 (0.349, 0.827) 0.00473 | 0.497 (0.253, 0.976) 0.0424 | 0.412 (0.21, 0.809) 0.01 |
| Intestinal disaccharidase deficiencies and disaccharide malabsorption (271.3) | endocrine/metabolic | 156/249/37 | 0.537 (0.349, 0.827) 0.00473 | 0.497 (0.253, 0.976) 0.0424 | 0.412 (0.21, 0.809) 0.01 |
| Disorders of phosphorus metabolism (275.53) | endocrine/metabolic | 35/30/70 | 4.3 (2.28, 8.12) 6.62e-06 | 6.46 (2.49, 16.7) 0.000122 | 2.87 (1.08, 7.57) 0.0337 |
| Disorders of fluid, electrolyte, and acid-base balance (276) | endocrine/metabolic | 21/3/173 | 2.18 (1.62, 2.93) 2.52e-07 | 3.47 (2.28, 5.29) 6.16e-09 | 1.47 (0.894, 2.42) 0.128 |
| Electrolyte imbalance (276.1) | endocrine/metabolic | 24/4/125 | 2.34 (1.68, 3.26) 5.55e-07 | 4.02 (2.48, 6.51) 1.5e-08 | 1.68 (0.959, 2.94) 0.0699 |
| Hyperosmolality and/or hypernatremia (276.11) | endocrine/metabolic | 41/160/103 | 20.5 (5.07, 83.2) 2.31e-05 | 66.8 (2.6, 1720) 0.0112 | 18.4 (0.925, 366) 0.0563 |
| Hyposmolality and/or hyponatremia (276.12) | endocrine/metabolic | 34/9/203 | 3.29 (1.97, 5.51) 5.86e-06 | 6.38 (3.09, 13.2) 5.66e-07 | 1.82 (0.78, 4.25) 0.166 |
| Hyperpotassemia (276.13) | endocrine/metabolic | 38/19/56 | 2.78 (1.76, 4.38) 1.02e-05 | 3.92 (2.09, 7.34) 1.99e-05 | 2.45 (1.12, 5.32) 0.0241 |
| Acid-base balance disorder (276.4) | endocrine/metabolic | 7/6/13 | 7.19 (3.91, 13.3) 2.42e-10 | 9.23 (3.94, 21.6) 3.03e-07 | 5.64 (2.08, 15.3) 0.000686 |
| Acidosis (276.41) | endocrine/metabolic | 11/11/22 | 6.65 (3.54, 12.5) 3.78e-09 | 7.77 (3.29, 18.4) 2.95e-06 | 4.91 (1.77, 13.6) 0.0022 |
| Hypovolemia (276.5) | endocrine/metabolic | 55/12/391 | 2.16 (1.47, 3.19) 9.75e-05 | 3.4 (2.02, 5.71) 3.89e-06 | 1.26 (0.677, 2.36) 0.463 |
| Fluid overload (276.6) | endocrine/metabolic | 33/96/24 | 5.83 (2.75, 12.4) 4.29e-06 | 5.04 (1.73, 14.7) 0.00304 | 5.11 (1.74, 14.9) 0.00292 |
| Immunity deficiency (279.1) | endocrine/metabolic | 134/48/546 | 2.84 (1.42, 5.7) 0.00323 | 6.41 (2.26, 18.2) 0.00047 | 1.2 (0.432, 3.33) 0.727 |
| Other anemias (285) | hematopoietic | 113/44/280 | 1.59 (1.19, 2.13) 0.00194 | 2.1 (1.4, 3.15) 0.000363 | 1.31 (0.809, 2.11) 0.274 |
| Acute posthemorrhagic anemia (285.1) | hematopoietic | 39/122/44 | 3.22 (1.91, 5.42) 1.08e-05 | 2.83 (1.36, 5.89) 0.00548 | 2.84 (1.26, 6.43) 0.0119 |
| Anemia of chronic disease (285.2) | hematopoietic | 22/21/30 | 3.98 (2.34, 6.75) 3.16e-07 | 4.15 (2.11, 8.15) 3.71e-05 | 3.81 (1.51, 9.66) 0.00472 |
| Anemia in chronic kidney disease (285.21) | hematopoietic | 124/303/35 | 2.82 (1.43, 5.56) 0.0027 | 2.27 (0.9, 5.72) 0.0825 | 5.04 (1.48, 17.2) 0.00975 |
| Diseases of white blood cells (288) | hematopoietic | 49/57/52 | 2.2 (1.5, 3.22) 5.81e-05 | 2.47 (1.46, 4.19) 0.000782 | 2.05 (1.11, 3.79) 0.0215 |
| Aphasia/speech disturbance (292.1) | mental disorders | 98/46/406 | 2.9 (1.52, 5.55) 0.00127 | 3.95 (1.83, 8.52) 0.000458 | 1.61 (0.408, 6.31) 0.498 |
| Epilepsy, recurrent seizures, convulsions (345) | neurological | 131/25/424 | 2.22 (1.31, 3.75) 0.00296 | 4.17 (2.07, 8.41) 6.39e-05 | 1.35 (0.514, 3.57) 0.539 |
| Epilepsy (345.1) | neurological | 223/27/NA | 2.75 (1.21, 6.26) 0.0159 | 11.2 (3.29, 38.3) 0.000114 |  |
| Convulsions (345.3) | neurological | 169/33/381 | 2.16 (1.25, 3.74) 0.0058 | 4 (1.92, 8.35) 0.000223 | 1.47 (0.543, 3.96) 0.451 |
| Hypermetropia (367.8) | sense organs | 96/352/48 | 0.398 (0.228, 0.697) 0.00126 | 0.554 (0.261, 1.18) 0.124 | 0.342 (0.139, 0.842) 0.0196 |
| Hypertension (401) | circulatory system | 26/51/87 | 2.07 (1.54, 2.78) 1.52e-06 | 2.08 (1.37, 3.14) 0.000522 | 1.69 (1.01, 2.85) 0.0473 |
| Essential hypertension (401.1) | circulatory system | 30/61/85 | 2.05 (1.52, 2.76) 2.53e-06 | 2.02 (1.33, 3.06) 0.000945 | 1.71 (1.01, 2.89) 0.0458 |
| Hypertensive heart and/or renal disease (401.2) | circulatory system | 9/14/12 | 4.6 (2.82, 7.52) 1.03e-09 | 4.62 (2.35, 9.11) 9.64e-06 | 4.06 (1.83, 9) 0.000553 |
| Hypertensive chronic kidney disease (401.22) | circulatory system | 25/40/16 | 4.53 (2.49, 8.23) 7.13e-07 | 4.78 (2.05, 11.1) 0.000287 | 4.79 (1.87, 12.3) 0.00109 |
| Other hypertensive complications (401.3) | circulatory system | 32/47/39 | 3.34 (2.01, 5.56) 3.57e-06 | 3.89 (1.82, 8.31) 0.000461 | 2.75 (1.27, 5.97) 0.0105 |
| Pulmonary heart disease (415) | circulatory system | 28/22/49 | 3.36 (2.05, 5.52) 1.62e-06 | 4.45 (2.18, 9.1) 4.27e-05 | 2.27 (1.14, 4.55) 0.0202 |
| Chronic pulmonary heart disease (415.2) | circulatory system | 36/49/31 | 4.36 (2.29, 8.3) 7.22e-06 | 6.51 (2.28, 18.6) 0.000475 | 3.14 (1.39, 7.08) 0.00575 |
| Abnormal electrocardiogram [ECG] [EKG] (426.7) | circulatory system | 86/36/320 | 1.74 (1.26, 2.42) 0.000829 | 2.37 (1.49, 3.76) 0.000258 | 1.31 (0.757, 2.26) 0.336 |
| Tachycardia NOS (427.7) | circulatory system | 29/10/94 | 2.55 (1.74, 3.74) 1.63e-06 | 3.5 (2.08, 5.89) 2.39e-06 | 1.87 (0.996, 3.53) 0.0515 |
| Congestive heart failure; nonhypertensive (428) | circulatory system | 43/45/147 | 2.51 (1.63, 3.85) 2.79e-05 | 2.94 (1.62, 5.34) 0.000392 | 1.83 (0.904, 3.71) 0.093 |
| Congestive heart failure (CHF) NOS (428.1) | circulatory system | 50/60/166 | 2.55 (1.61, 4.03) 6.38e-05 | 2.85 (1.53, 5.32) 0.000944 | 1.84 (0.857, 3.96) 0.118 |
| Abnormal function study of cardiovascular system (429.2) | circulatory system | 90/41/600 | 1.78 (1.26, 2.5) 0.000952 | 2.45 (1.51, 3.98) 0.00029 | 1.06 (0.603, 1.88) 0.831 |
| Peripheral vascular disease (443) | circulatory system | 132/38/576 | 2.36 (1.34, 4.17) 0.00307 | 3.74 (1.83, 7.62) 0.000284 | 1.18 (0.391, 3.55) 0.771 |
| Other specified peripheral vascular diseases (443.8) | circulatory system | 117/37/656 | 2.71 (1.43, 5.14) 0.00231 | 4.56 (2.02, 10.3) 0.000261 | 0.95 (0.297, 3.04) 0.931 |
| Peripheral vascular disease, unspecified (443.9) | circulatory system | 69/26/616 | 3.87 (1.88, 7.96) 0.000238 | 6.53 (2.58, 16.5) 7.51e-05 | 1.11 (0.328, 3.77) 0.865 |
| Hypotension (458) | circulatory system | 15/17/14 | 3.4 (2.22, 5.21) 2.04e-08 | 3.44 (1.96, 6.04) 1.69e-05 | 4.12 (1.81, 9.4) 0.000753 |
| Iatrogenic hypotension (458.2) | circulatory system | 149/387/47 | 4.69 (1.63, 13.5) 0.00424 | 2.55 (0.696, 9.31) 0.158 | 12 (1.52, 94.8) 0.0185 |
| Hypotension NOS (458.9) | circulatory system | 19/23/27 | 3.3 (2.11, 5.15) 1.45e-07 | 3.34 (1.86, 6) 5.4e-05 | 3.56 (1.53, 8.3) 0.00324 |
| Other disorders of circulatory system (459) | circulatory system | 37/35/34 | 2.34 (1.61, 3.4) 8.69e-06 | 2.6 (1.56, 4.33) 0.000241 | 2.26 (1.26, 4.05) 0.00635 |
| Acute upper respiratory infections of multiple or unspecified sites (465) | respiratory | 94/211/43 | 0.627 (0.473, 0.83) 0.00111 | 0.645 (0.439, 0.948) 0.0255 | 0.537 (0.332, 0.87) 0.0116 |
| Allergic rhinitis (476) | respiratory | 67/351/41 | 0.547 (0.397, 0.754) 0.000228 | 0.717 (0.471, 1.09) 0.122 | 0.478 (0.271, 0.843) 0.0107 |
| Pneumonia (480) | respiratory | 20/24/65 | 2.28 (1.67, 3.11) 1.79e-07 | 2.45 (1.59, 3.79) 5.48e-05 | 1.79 (1.06, 3.04) 0.0308 |
| Viral pneumonia (480.2) | respiratory | 6/28/5 | 7.46 (4.13, 13.5) 2.56e-11 | 5.33 (2.28, 12.5) 0.000116 | 7.97 (2.75, 23.1) 0.000129 |
| Pulmonary congestion and hypostasis (503) | respiratory | 48/129/38 | 8.35 (2.98, 23.4) 5.33e-05 | 5.41 (1.61, 18.1) 0.0062 | 12 (1.81, 80) 0.0101 |
| Other pulmonary inflamation or edema (505) | respiratory | 27/34/25 | 7.08 (3.18, 15.7) 1.58e-06 | 8.82 (2.77, 28.1) 0.00023 | 5.87 (1.83, 18.8) 0.00293 |
| Empyema and pneumothorax (506) | respiratory | 23/93/10 | 4.55 (2.54, 8.14) 3.3e-07 | 3.36 (1.52, 7.41) 0.00273 | 6.81 (2.46, 18.9) 0.000226 |
| Pleurisy; pleural effusion (507) | respiratory | 16/15/33 | 4.43 (2.62, 7.5) 2.99e-08 | 5.66 (2.61, 12.3) 1.11e-05 | 3.07 (1.38, 6.81) 0.00579 |
| Pulmonary collapse; interstitial and compensatory emphysema (508) | respiratory | 18/7/218 | 5.97 (3.14, 11.3) 4.72e-08 | 11.8 (4.57, 30.4) 3.36e-07 | 1.94 (0.733, 5.11) 0.182 |
| Respiratory failure, insufficiency, arrest (509) | respiratory | 1/2/1 | 6.78 (4.51, 10.2) 4.3e-20 | 5.45 (3.09, 9.64) 5.26e-09 | 7.1 (3.56, 14.1) 2.62e-08 |
| Respiratory failure (509.1) | respiratory | 2/8/2 | 8.64 (5.24, 14.2) 2.62e-17 | 5.45 (2.83, 10.5) 3.98e-07 | 10.8 (4.29, 27.2) 4.38e-07 |
| Respiratory insufficiency (509.2) | respiratory | 5/13/17 | 7.05 (3.98, 12.5) 2.14e-11 | 7 (3.03, 16.1) 5.08e-06 | 4.26 (1.72, 10.5) 0.00171 |
| Pulmonary insufficiency or respiratory failure following trauma and surgery (509.3) | respiratory | 10/50/3 | 14.5 (5.99, 34.9) 2.79e-09 | 12.4 (3.02, 50.6) 0.000477 | 19.1 (4.42, 82.6) 7.8e-05 |
| Dependence on respirator [Ventilator] or supplemental oxygen (509.8) | respiratory | 8/18/6 | 8.15 (4.2, 15.8) 6.01e-10 | 7.95 (3.09, 20.5) 1.74e-05 | 8.23 (2.79, 24.2) 0.000133 |
| Other diseases of lung (510) | respiratory | 270/42/327 | 1.55 (1.03, 2.32) 0.0336 | 2.76 (1.58, 4.82) 0.000343 | 0.728 (0.375, 1.41) 0.348 |
| Cough (512.8) | respiratory | 766/452/40 | 0.89 (0.657, 1.21) 0.451 | 1.29 (0.854, 1.94) 0.228 | 0.489 (0.282, 0.846) 0.0106 |
| Dysphagia (532) | digestive | 187/39/657 | 1.7 (1.14, 2.53) 0.00864 | 2.68 (1.57, 4.57) 0.000285 | 0.972 (0.49, 1.93) 0.936 |
| Renal failure (585) | genitourinary | 4/5/4 | 3.32 (2.36, 4.66) 4.55e-12 | 3.67 (2.28, 5.92) 8.95e-08 | 3.11 (1.76, 5.48) 9e-05 |
| Acute renal failure (585.1) | genitourinary | 3/1/9 | 4.33 (2.9, 6.45) 6.06e-13 | 5.42 (3.08, 9.57) 5.22e-09 | 3.44 (1.79, 6.59) 0.000202 |
| Chronic renal failure [CKD] (585.3) | genitourinary | 13/20/7 | 3.08 (2.11, 4.49) 5.82e-09 | 3.13 (1.85, 5.28) 1.99e-05 | 3.42 (1.81, 6.46) 0.000147 |
| End stage renal disease (585.32) | genitourinary | 57/73/50 | 3.67 (1.9, 7.06) 0.000102 | 6.83 (2.15, 21.8) 0.00114 | 2.83 (1.18, 6.83) 0.0204 |
| Chronic Kidney Disease, Stage III (585.33) | genitourinary | 31/29/23 | 3.26 (1.99, 5.34) 2.81e-06 | 3.73 (1.91, 7.3) 0.000121 | 3.6 (1.58, 8.19) 0.00222 |
| Chronic Kidney Disease, Stage IV (585.34) | genitourinary | 53/126/28 | 3.99 (2.01, 7.93) 8.01e-05 | 3.92 (1.48, 10.4) 0.00602 | 5.34 (1.73, 16.4) 0.00351 |
| Chronic kidney disease, Stage I or II (585.4) | genitourinary | 42/226/11 | 3.98 (2.09, 7.55) 2.45e-05 | 2.82 (1.09, 7.32) 0.0329 | 6.33 (2.3, 17.4) 0.000358 |
| Other disorders of the kidney and ureters (586) | genitourinary | 45/16/90 | 2.12 (1.48, 3.04) 4.08e-05 | 2.97 (1.81, 4.86) 1.53e-05 | 1.83 (1, 3.33) 0.0488 |

| Trait (Phecode) | Category | Rank All/NHW/NHB | All OR (95% CI) P-value | NHW OR (95% CI) P-value | NHB OR (95% CI) P-value |
| --- | --- | --- | --- | --- | --- |
| Septicemia (038) | infectious diseases | 8/38/12 | 4.73 (3, 7.47) 2.31e-11 | 3.74 (1.94, 7.2) 7.83e-05 | 4.58 (2.12, 9.9) 0.000107 |
| Bacteremia (038.3) | infectious diseases | 17/31/13 | 8.91 (4.27, 18.6) 5.79e-09 | 7.85 (2.89, 21.3) 5.21e-05 | 10.3 (3.07, 34.9) 0.000163 |
| Staphylococcus infections (041.1) | infectious diseases | 103/20/662 | 3.52 (1.76, 7.04) 0.000368 | 7.38 (3, 18.2) 1.41e-05 | 0.93 (0.199, 4.34) 0.926 |
| Methicillin sensitive Staphylococcus aureus (041.11) | infectious diseases | 101/32/NA | 4.6 (1.99, 10.6) 0.000361 | 9.69 (3.22, 29.2) 5.37e-05 |  |
| Streptococcus infection (041.2) | infectious diseases | 56/98/49 | 4.44 (2.19, 9.03) 3.79e-05 | 4.9 (1.88, 12.8) 0.00117 | 5.64 (1.69, 18.9) 0.005 |
| Protein-calorie malnutrition (260) | endocrine/metabolic | 62/11/270 | 2.94 (1.75, 4.96) 5.13e-05 | 5.37 (2.7, 10.7) 1.59e-06 | 1.72 (0.69, 4.3) 0.243 |
| Proteinuria (269) | endocrine/metabolic | 337/606/50 | 1.71 (1.01, 2.88) 0.0459 | 1.36 (0.613, 3) 0.452 | 2.96 (1.38, 6.37) 0.00538 |
| Disorders of mineral metabolism (275) | endocrine/metabolic | 28/77/30 | 2.85 (1.9, 4.27) 4.34e-07 | 2.67 (1.53, 4.67) 0.000541 | 2.9 (1.49, 5.65) 0.00171 |
| Disorders of calcium/phosphorus metabolism (275.5) | endocrine/metabolic | 39/69/106 | 2.77 (1.78, 4.31) 6.02e-06 | 2.98 (1.63, 5.43) 0.000372 | 2.29 (1.08, 4.84) 0.0302 |
| Disorders of phosphorus metabolism (275.53) | endocrine/metabolic | 31/26/78 | 4.08 (2.31, 7.22) 1.33e-06 | 5.85 (2.51, 13.6) 4.2e-05 | 2.86 (1.21, 6.77) 0.017 |
| Disorders of fluid, electrolyte, and acid-base balance (276) | endocrine/metabolic | 21/2/101 | 2.51 (1.8, 3.48) 4.39e-08 | 3.65 (2.28, 5.83) 6.26e-08 | 1.84 (1.06, 3.17) 0.0292 |
| Electrolyte imbalance (276.1) | endocrine/metabolic | 22/1/122 | 2.69 (1.88, 3.85) 6.75e-08 | 4.36 (2.6, 7.32) 2.61e-08 | 1.84 (1.01, 3.35) 0.0451 |
| Hyperosmolality and/or hypernatremia (276.11) | endocrine/metabolic | 20/157/29 | 11.1 (4.7, 26.4) 4.23e-08 | 7.05 (1.78, 27.9) 0.00538 | 19.5 (3.11, 123) 0.00153 |
| Hyposmolality and/or hyponatremia (276.12) | endocrine/metabolic | 82/7/486 | 2.7 (1.62, 4.5) 0.000147 | 6 (3, 12) 4.06e-07 | 1.26 (0.523, 3.03) 0.607 |
| Hyperpotassemia (276.13) | endocrine/metabolic | 54/62/46 | 2.67 (1.68, 4.23) 3.05e-05 | 3.43 (1.78, 6.62) 0.000232 | 2.95 (1.4, 6.21) 0.00443 |
| Acid-base balance disorder (276.4) | endocrine/metabolic | 4/4/4 | 7.53 (4.46, 12.7) 3.93e-14 | 7.69 (3.63, 16.3) 9.8e-08 | 7.06 (3.06, 16.3) 4.59e-06 |
| Acidosis (276.41) | endocrine/metabolic | 6/10/8 | 7.47 (4.32, 12.9) 6.14e-13 | 7.1 (3.26, 15.5) 8.26e-07 | 6.72 (2.81, 16.1) 1.9e-05 |
| Hypovolemia (276.5) | endocrine/metabolic | 77/28/182 | 2.28 (1.5, 3.47) 0.000123 | 3.24 (1.84, 5.69) 4.4e-05 | 1.77 (0.89, 3.53) 0.104 |
| Fluid overload (276.6) | endocrine/metabolic | 25/67/19 | 5.35 (2.82, 10.2) 2.78e-07 | 6.17 (2.27, 16.7) 0.000354 | 4.95 (2.02, 12.2) 0.000488 |
| Immunity deficiency (279.1) | endocrine/metabolic | 73/40/183 | 3.91 (1.97, 7.75) 9.19e-05 | 7.29 (2.69, 19.8) 9.45e-05 | 2.34 (0.84, 6.5) 0.104 |
| Aplastic anemia (284) | hematopoietic | 141/50/NA | 4.5 (1.75, 11.6) 0.00183 | 7.83 (2.7, 22.8) 0.000156 |  |
| Other anemias (285) | hematopoietic | 58/16/333 | 2.02 (1.44, 2.83) 4.09e-05 | 3.02 (1.89, 4.82) 3.56e-06 | 1.3 (0.753, 2.24) 0.348 |
| Acute posthemorrhagic anemia (285.1) | hematopoietic | 45/52/148 | 3.22 (1.92, 5.42) 1.02e-05 | 4.3 (2.01, 9.18) 0.000166 | 2.13 (0.95, 4.78) 0.0663 |
| Anemia of chronic disease (285.2) | hematopoietic | 30/37/48 | 3.6 (2.16, 5.98) 8.33e-07 | 4.02 (2.02, 8) 7.57e-05 | 3.29 (1.44, 7.49) 0.00465 |
| Anemia in neoplastic disease (285.22) | hematopoietic | 76/49/NA | 9.58 (3.03, 30.3) 0.000119 | 15.2 (3.73, 61.5) 0.000144 |  |
| Encounter for long-term (current) use of anticoagulants (286.2) | hematopoietic | 49/121/34 | 3.08 (1.85, 5.14) 1.65e-05 | 3.08 (1.49, 6.37) 0.00246 | 3.39 (1.54, 7.46) 0.00239 |
| Purpura and other hemorrhagic conditions (287) | hematopoietic | 63/27/302 | 2.75 (1.68, 4.51) 5.5e-05 | 3.74 (1.99, 7.02) 4.25e-05 | 1.62 (0.658, 4) 0.294 |
| Thrombocytopenia (287.3) | hematopoietic | 51/19/289 | 3.06 (1.84, 5.1) 1.68e-05 | 4.64 (2.37, 9.08) 7.62e-06 | 1.66 (0.67, 4.13) 0.272 |
| Secondary thrombocytopenia (287.32) | hematopoietic | 48/NA/NA | 18.4 (4.94, 68.3) 1.41e-05 |  |  |
| Diseases of white blood cells (288) | hematopoietic | 46/13/248 | 2.46 (1.65, 3.68) 1.13e-05 | 3.78 (2.18, 6.57) 2.23e-06 | 1.51 (0.792, 2.88) 0.211 |
| Decreased white blood cell count (288.1) | hematopoietic | 238/44/524 | 2.14 (1.16, 3.94) 0.0143 | 5.67 (2.36, 13.6) 0.000101 | 0.797 (0.292, 2.17) 0.658 |
| Neutropenia (288.11) | hematopoietic | 108/30/477 | 4.18 (1.88, 9.27) 0.000436 | 8.5 (3.03, 23.8) 4.76e-05 | 1.42 (0.397, 5.11) 0.588 |
| Elevated white blood cell count (288.2) | hematopoietic | 66/22/245 | 2.4 (1.57, 3.67) 5.68e-05 | 3.68 (2.02, 6.69) 1.96e-05 | 1.55 (0.79, 3.03) 0.204 |
| Delirium due to conditions classified elsewhere (290.2) | mental disorders | 70/428/21 | 4.18 (2.05, 8.51) 8.04e-05 | 1.86 (0.726, 4.76) 0.196 | 27.2 (4.13, 179) 0.000589 |
| Chronic pain syndrome (355.1) | neurological | 356/707/41 | 2.49 (0.971, 6.4) 0.0575 | 0.611 (0.0985, 3.79) 0.597 | 9.2 (2.09, 40.6) 0.00338 |
| Hypertensive heart and/or renal disease (401.2) | circulatory system | 27/21/45 | 3.94 (2.32, 6.69) 3.63e-07 | 5.11 (2.44, 10.7) 1.53e-05 | 3.51 (1.49, 8.27) 0.00403 |
| Hypertensive chronic kidney disease (401.22) | circulatory system | 34/58/42 | 4.39 (2.39, 8.04) 1.75e-06 | 5.22 (2.19, 12.5) 0.000201 | 4.04 (1.58, 10.3) 0.00356 |
| Ischemic Heart Disease (411) | circulatory system | 36/34/40 | 2.45 (1.68, 3.59) 3.55e-06 | 2.96 (1.74, 5.03) 5.96e-05 | 2.55 (1.37, 4.77) 0.0033 |
| Myocardial infarction (411.2) | circulatory system | 55/42/90 | 3.03 (1.79, 5.12) 3.43e-05 | 3.78 (1.94, 7.38) 9.55e-05 | 3.16 (1.18, 8.46) 0.0224 |
| Coronary atherosclerosis (411.4) | circulatory system | 40/76/17 | 2.56 (1.7, 3.86) 7.07e-06 | 2.79 (1.56, 4.98) 0.000526 | 3.34 (1.73, 6.43) 0.000316 |
| Other forms of chronic heart disease (414) | circulatory system | 84/215/24 | 2.89 (1.66, 5.01) 0.000161 | 2.57 (1.15, 5.72) 0.021 | 4.02 (1.77, 9.16) 0.000925 |
| Pulmonary heart disease (415) | circulatory system | 29/36/38 | 3.37 (2.1, 5.41) 5.01e-07 | 3.98 (2.01, 7.89) 7.45e-05 | 2.76 (1.41, 5.42) 0.00306 |
| Chronic pulmonary heart disease (415.2) | circulatory system | 44/75/36 | 3.58 (2.04, 6.3) 9.52e-06 | 4.99 (2.01, 12.4) 0.000521 | 3.11 (1.49, 6.48) 0.00246 |
| Cardiac conduction disorders (426) | circulatory system | 90/29/253 | 1.99 (1.38, 2.86) 0.000222 | 3.04 (1.78, 5.19) 4.72e-05 | 1.45 (0.806, 2.61) 0.215 |
| Abnormal electrocardiogram [ECG] [EKG] (426.7) | circulatory system | 71/24/211 | 2.1 (1.45, 3.05) 8.14e-05 | 3.24 (1.87, 5.6) 2.56e-05 | 1.54 (0.846, 2.79) 0.158 |
| Other cardiac conduction disorders (426.8) | circulatory system | 60/43/170 | 4.72 (2.24, 9.95) 4.47e-05 | 6.64 (2.56, 17.2) 9.92e-05 | 2.9 (0.835, 10.1) 0.0937 |
| Cardiac dysrhythmias (427) | circulatory system | 125/47/359 | 1.76 (1.26, 2.45) 0.000874 | 2.56 (1.58, 4.15) 0.000132 | 1.27 (0.734, 2.19) 0.395 |
| Tachycardia NOS (427.7) | circulatory system | 42/15/124 | 2.58 (1.7, 3.91) 7.75e-06 | 3.99 (2.23, 7.14) 3.19e-06 | 1.97 (1.01, 3.84) 0.0459 |
| Congestive heart failure; nonhypertensive (428) | circulatory system | 53/39/81 | 2.58 (1.66, 4.01) 2.57e-05 | 3.66 (1.91, 6.99) 8.79e-05 | 2.3 (1.15, 4.56) 0.0178 |
| Abnormal function study of cardiovascular system (429.2) | circulatory system | 106/33/416 | 1.98 (1.36, 2.9) 0.000406 | 3.07 (1.78, 5.31) 5.84e-05 | 1.24 (0.665, 2.32) 0.498 |
| Hypotension (458) | circulatory system | 7/5/10 | 4.22 (2.78, 6.42) 1.49e-11 | 4.41 (2.49, 7.81) 3.6e-07 | 4.75 (2.25, 10) 4.38e-05 |
| Iatrogenic hypotension (458.2) | circulatory system | 116/432/26 | 4.71 (1.92, 11.5) 0.000694 | 2.34 (0.635, 8.64) 0.201 | 11.8 (2.67, 52.5) 0.00115 |
| Hypotension NOS (458.9) | circulatory system | 14/12/20 | 4.01 (2.59, 6.2) 5.19e-10 | 4.28 (2.36, 7.75) 1.66e-06 | 4.04 (1.83, 8.91) 0.000555 |
| Other disorders of circulatory system (459) | circulatory system | 23/59/11 | 2.99 (2, 4.46) 7.95e-08 | 2.88 (1.65, 5.03) 0.000202 | 3.61 (1.94, 6.73) 5.39e-05 |
| Circulatory disease NEC (459.9) | circulatory system | 38/130/18 | 2.63 (1.73, 3.99) 5.81e-06 | 2.44 (1.35, 4.41) 0.00302 | 3.21 (1.68, 6.12) 0.000401 |
| Bacterial pneumonia (480.1) | respiratory | 172/451/44 | 2.56 (1.35, 4.87) 0.00403 | 1.82 (0.695, 4.75) 0.224 | 5.06 (1.69, 15.1) 0.00375 |
| Chronic airway obstruction (496) | respiratory | 50/25/108 | 2.83 (1.76, 4.54) 1.67e-05 | 3.8 (2.02, 7.15) 3.34e-05 | 2.42 (1.08, 5.45) 0.0326 |
| Pulmonary congestion and hypostasis (503) | respiratory | 59/145/39 | 5.77 (2.49, 13.4) 4.36e-05 | 5.12 (1.67, 15.7) 0.00431 | 7.72 (1.99, 29.9) 0.00309 |
| Other pulmonary inflammation or edema (505) | respiratory | 26/133/7 | 6.21 (3.09, 12.5) 2.78e-07 | 4.61 (1.67, 12.7) 0.00309 | 11.1 (3.68, 33.3) 1.86e-05 |
| Empyema and pneumothorax (506) | respiratory | 15/48/9 | 5.62 (3.23, 9.77) 1.02e-09 | 4.77 (2.14, 10.7) 0.000135 | 6.37 (2.64, 15.4) 3.86e-05 |
| Pleurisy; pleural effusion (507) | respiratory | 33/72/57 | 3.64 (2.15, 6.19) 1.7e-06 | 3.93 (1.84, 8.39) 0.000405 | 3.18 (1.38, 7.34) 0.00664 |
| Pulmonary collapse; interstitial and compensatory emphysema (508) | respiratory | 16/3/114 | 6.05 (3.36, 10.9) 2.06e-09 | 9.08 (4.04, 20.4) 8.98e-08 | 2.91 (1.06, 8.03) 0.0387 |
| Respiratory failure, insufficiency, arrest (509) | respiratory | 1/23/1 | 4.86 (3.3, 7.15) 1.04e-15 | 3.48 (1.96, 6.19) 2.23e-05 | 6.09 (3.26, 11.4) 1.5e-08 |
| Respiratory failure (509.1) | respiratory | 5/90/2 | 5.07 (3.27, 7.84) 3.45e-13 | 3.08 (1.59, 5.95) 0.000853 | 6.79 (3.31, 13.9) 1.72e-07 |
| Respiratory insufficiency (509.2) | respiratory | 2/14/5 | 7.92 (4.76, 13.2) 1.94e-15 | 6.12 (2.88, 13) 2.57e-06 | 6.6 (2.8, 15.5) 1.6e-05 |
| Pulmonary insufficiency or respiratory failure following trauma and surgery (509.3) | respiratory | 3/17/3 | 18.2 (8.79, 37.7) 5.65e-15 | 18.1 (5.17, 63.3) 5.85e-06 | 16.6 (5.73, 47.9) 2.2e-07 |
| Dependence on respirator [Ventilator] or supplemental oxygen (509.8) | respiratory | 11/74/6 | 6.68 (3.74, 11.9) 1.36e-10 | 4.57 (1.96, 10.7) 0.000453 | 7.99 (3.1, 20.6) 1.73e-05 |
| Renal failure (585) | genitourinary | 9/8/16 | 3.39 (2.36, 4.89) 5.33e-11 | 3.81 (2.26, 6.41) 4.92e-07 | 3.07 (1.67, 5.62) 0.000289 |
| Acute renal failure (585.1) | genitourinary | 10/9/25 | 3.78 (2.52, 5.65) 1.02e-10 | 4.38 (2.46, 7.81) 5.63e-07 | 3.04 (1.56, 5.91) 0.00104 |
| Chronic renal failure [CKD] (585.3) | genitourinary | 18/18/15 | 3.27 (2.19, 4.88) 7.38e-09 | 3.71 (2.1, 6.56) 6.24e-06 | 3.39 (1.76, 6.54) 0.000272 |
| Renal dialysis (585.31) | genitourinary | 41/119/66 | 4.54 (2.34, 8.81) 7.5e-06 | 5.37 (1.83, 15.7) 0.00216 | 3.46 (1.32, 9.1) 0.0118 |

| Trait (Phecode) | Category | Rank All/NHW/NHB | All OR (95% CI) P-value | NHW OR (95% CI) P-value | NHB OR (95% CI) P-value |
| --- | --- | --- | --- | --- | --- |
| Septicemia (038) | infectious diseases | 10/114/26 | 4.31 (2.41, 7.72) 9.03e-07 | 2.73 (1.05, 7.14) 0.0403 | 4.41 (1.74, 11.2) 0.00172 |
| Staphylococcus infections (041.1) | infectious diseases | 215/25/546 | 2.92 (1.09, 7.84) 0.0335 | 7.51 (2.1, 26.9) 0.00194 | 0.602 (0.0299, 12.1) 0.74 |
| Methicillin sensitive Staphylococcus aureus (041.11) | infectious diseases | 179/38/NA | 3.82 (1.24, 11.7) 0.0193 | 9.09 (2.1, 39.3) 0.00316 |  |
| Streptococcus infection (041.2) | infectious diseases | 212/41/398 | 3.1 (1.1, 8.7) 0.0321 | 6.88 (1.9, 24.9) 0.00328 | 1.98 (0.304, 12.9) 0.475 |
| Secondary diabetes mellitus (249) | endocrine/metabolic | 92/12/242 | 4.4 (1.75, 11) 0.00164 | 6.9 (2.25, 21.2) 0.000739 | 2.81 (0.511, 15.4) 0.235 |
| Type 2 diabetes with neurological manifestations (250.24) | endocrine/metabolic | 146/366/30 | 2.78 (1.33, 5.82) 0.00663 | 1.88 (0.65, 5.44) 0.244 | 6.52 (1.99, 21.4) 0.00199 |
| Diabetes type 2 with peripheral circulatory disorders (250.25) | endocrine/metabolic | 15/19/18 | 7.38 (3.13, 17.4) 4.99e-06 | 6.21 (2.06, 18.7) 0.00117 | 12 (2.73, 53) 0.00102 |
| Disorders of mineral metabolism (275) | endocrine/metabolic | 46/284/19 | 2.8 (1.6, 4.91) 0.000301 | 1.84 (0.781, 4.33) 0.163 | 4.21 (1.77, 9.99) 0.00112 |
| Disorders of phosphorus metabolism (275.53) | endocrine/metabolic | 32/78/97 | 4.06 (1.95, 8.47) 0.000183 | 4.13 (1.29, 13.2) 0.0171 | 3.26 (1.09, 9.73) 0.034 |
| Disorders of fluid, electrolyte, and acid-base balance (276) | endocrine/metabolic | 13/104/8 | 3.28 (1.99, 5.41) 3.24e-06 | 2.29 (1.08, 4.87) 0.0306 | 5.98 (2.38, 15.1) 0.000148 |
| Electrolyte imbalance (276.1) | endocrine/metabolic | 16/64/12 | 3.36 (1.99, 5.66) 5.45e-06 | 2.84 (1.3, 6.2) 0.00879 | 5.41 (2.08, 14.1) 0.000555 |
| Hyperosmolality and/or hypernatremia (276.11) | endocrine/metabolic | 4/26/3 | 11.5 (4.86, 27.1) 2.56e-08 | 11.3 (2.43, 52.6) 0.00198 | 19.1 (4.56, 79.7) 5.42e-05 |
| Hyposmolality and/or hyponatremia (276.12) | endocrine/metabolic | 28/13/93 | 4.03 (1.99, 8.14) 0.000106 | 4.95 (1.94, 12.6) 0.000804 | 3.89 (1.12, 13.5) 0.0322 |
| Hyperpotassemia (276.13) | endocrine/metabolic | 39/116/31 | 3.4 (1.77, 6.51) 0.000226 | 2.68 (1.04, 6.85) 0.0404 | 5.68 (1.86, 17.4) 0.00229 |
| Hypopotassemia (276.14) | endocrine/metabolic | 21/127/32 | 4.63 (2.25, 9.55) 3.28e-05 | 3.07 (1.01, 9.31) 0.0473 | 5.96 (1.88, 18.9) 0.00239 |
| Acid-base balance disorder (276.4) | endocrine/metabolic | 2/45/7 | 7.05 (3.63, 13.7) 8.14e-09 | 4.33 (1.58, 11.9) 0.00445 | 10.6 (3.29, 34.2) 7.79e-05 |
| Acidosis (276.41) | endocrine/metabolic | 7/67/11 | 6.79 (3.38, 13.6) 7.38e-08 | 4.02 (1.41, 11.5) 0.00948 | 8.71 (2.55, 29.7) 0.000548 |
| Alkalosis (276.42) | endocrine/metabolic | 20/NA/NA | 19.1 (4.77, 76.9) 3.17e-05 |  |  |
| Hypovolemia (276.5) | endocrine/metabolic | 84/399/40 | 3.02 (1.55, 5.88) 0.00119 | 1.68 (0.657, 4.31) 0.278 | 5.43 (1.78, 16.6) 0.00299 |
| Fluid overload (276.6) | endocrine/metabolic | 26/93/67 | 5.87 (2.44, 14.1) 7.91e-05 | 4.21 (1.21, 14.7) 0.0243 | 5.58 (1.45, 21.4) 0.0123 |
| Aplastic anemia (284) | hematopoietic | 173/15/NA | 5.33 (1.35, 21.1) 0.0172 | 12.5 (2.78, 55.8) 0.000976 |  |
| Pancytopenia (284.1) | hematopoietic | 174/16/NA | 5.33 (1.35, 21.1) 0.0172 | 12.5 (2.78, 55.8) 0.000976 |  |
| Encounter for long-term (current) use of anticoagulants (286.2) | hematopoietic | 190/701/47 | 2.32 (1.12, 4.79) 0.023 | 1.29 (0.406, 4.07) 0.669 | 4.55 (1.57, 13.2) 0.00517 |
| Diseases of white blood cells (288) | hematopoietic | 45/2/126 | 2.86 (1.62, 5.06) 0.000299 | 4.96 (2.19, 11.3) 0.000128 | 2.26 (0.938, 5.45) 0.069 |
| Decreased white blood cell count (288.1) | hematopoietic | 560/28/612 | 1.58 (0.605, 4.15) 0.348 | 8.38 (2.17, 32.4) 0.00206 | 0.869 (0.197, 3.84) 0.854 |
| Elevated white blood cell count (288.2) | hematopoietic | 27/1/119 | 3.19 (1.78, 5.73) 0.000101 | 5.86 (2.54, 13.6) 3.51e-05 | 2.39 (0.951, 5.99) 0.0638 |
| Delirium dementia and amnestic and other cognitive disorders (290) | mental disorders | 25/73/10 | 4.48 (2.15, 9.34) 6.21e-05 | 3.53 (1.28, 9.75) 0.0151 | 8.69 (2.65, 28.5) 0.00036 |
| Delirium due to conditions classified elsewhere (290.2) | mental disorders | 11/69/6 | 8.55 (3.63, 20.2) 9.35e-07 | 4.55 (1.4, 14.8) 0.0117 | 15.3 (3.99, 58.4) 6.93e-05 |
| Other persistent mental disorders due to conditions classified elsewhere (290.3) | mental disorders | 43/58/64 | 4.65 (2.03, 10.6) 0.000283 | 4.72 (1.52, 14.7) 0.0074 | 6.59 (1.56, 27.9) 0.0104 |
| Alteration of consciousness (291.8) | mental disorders | 100/131/49 | 3.24 (1.54, 6.82) 0.00199 | 2.87 (1, 8.22) 0.0493 | 4.97 (1.61, 15.4) 0.00533 |
| Neurological disorders (292) | mental disorders | 83/8/152 | 2.52 (1.44, 4.4) 0.00117 | 4.41 (1.93, 10.1) 0.000438 | 2.17 (0.869, 5.42) 0.097 |
| Aphasia/speech disturbance (292.1) | mental disorders | 72/6/471 | 4.44 (1.86, 10.6) 0.000806 | 7.05 (2.51, 19.8) 0.000212 | 1.69 (0.225, 12.6) 0.611 |
| Aphasia (292.11) | mental disorders | 62/9/NA | 8.47 (2.53, 28.4) 0.000534 | 11.2 (2.9, 43.1) 0.000457 |  |
| Mild cognitive impairment (292.2) | mental disorders | 91/21/NA | 5.79 (1.95, 17.2) 0.00158 | 9.21 (2.38, 35.7) 0.00132 |  |
| Memory loss (292.3) | mental disorders | 54/18/361 | 4.06 (1.87, 8.83) 0.000397 | 5.2 (1.94, 14) 0.00107 | 2.03 (0.399, 10.3) 0.394 |
| Altered mental status (292.4) | mental disorders | 85/35/52 | 2.83 (1.5, 5.32) 0.00128 | 4.02 (1.61, 10) 0.00285 | 3.82 (1.45, 10.1) 0.00655 |
| Alcoholic liver damage (317.11) | mental disorders | 36/NA/NA | 13.7 (3.45, 54.5) 0.000202 |  |  |
| Epilepsy, recurrent seizures, convulsions (345) | neurological | 263/605/42 | 2.26 (0.962, 5.33) 0.0614 | 1.48 (0.403, 5.44) 0.555 | 7.24 (1.95, 26.9) 0.00311 |
| Convulsions (345.3) | neurological | 274/679/41 | 2.33 (0.942, 5.76) 0.0673 | 1.45 (0.323, 6.49) 0.629 | 7.39 (1.97, 27.7) 0.00302 |
| Other conditions of brain (348) | neurological | 74/137/20 | 3.27 (1.63, 6.58) 0.000868 | 2.68 (0.985, 7.3) 0.0536 | 5.83 (2.02, 16.9) 0.00113 |
| Retinal vascular changes and abnomalities (362.4) | sense organs | 241/36/NA | 3.13 (1.02, 9.59) 0.046 | 9.59 (2.17, 42.4) 0.00287 |  |
| Blindness and low vision (367.9) | sense organs | 159/34/584 | 3.31 (1.3, 8.42) 0.0119 | 6.31 (1.9, 21) 0.00266 | 1.27 (0.176, 9.19) 0.811 |
| Pulmonary heart disease (415) | circulatory system | 40/52/101 | 3.33 (1.75, 6.33) 0.000237 | 3.75 (1.46, 9.65) 0.00612 | 2.63 (1.05, 6.56) 0.0385 |
| Cardiac conduction disorders (426) | circulatory system | 77/166/45 | 2.62 (1.48, 4.64) 0.000943 | 2.16 (0.932, 4.99) 0.0727 | 4.5 (1.63, 12.4) 0.00371 |
| Atrioventricular [AV] block (426.2) | circulatory system | 47/180/39 | 5.04 (2.1, 12.1) 0.000302 | 3.23 (0.857, 12.2) 0.0832 | 7.49 (2, 28.1) 0.00284 |
| First degree AV block (426.21) | circulatory system | 24/149/15 | 7.25 (2.8, 18.8) 4.54e-05 | 3.71 (0.946, 14.5) 0.06 | 122 (7.53, 1980) 0.000723 |
| Abnormal electrocardiogram [ECG] [EKG] (426.7) | circulatory system | 68/143/48 | 2.71 (1.52, 4.85) 0.000739 | 2.27 (0.973, 5.3) 0.058 | 4.31 (1.55, 12) 0.00518 |
| Other cardiac conduction disorders (426.8) | circulatory system | 107/49/466 | 5.15 (1.79, 14.9) 0.0024 | 6.32 (1.75, 22.9) 0.00498 | 1.77 (0.204, 15.4) 0.604 |
| Cardiac defibrillator in situ (426.92) | circulatory system | 150/NA/43 | 4.55 (1.49, 14) 0.00795 |  | 7.84 (1.97, 31.2) 0.00351 |
| Cardiac dysrhythmias (427) | circulatory system | 115/328/37 | 2.29 (1.33, 3.93) 0.00274 | 1.69 (0.739, 3.87) 0.214 | 4.64 (1.71, 12.6) 0.00258 |
| Atrial fibrillation and flutter (427.2) | circulatory system | 57/367/5 | 3.4 (1.72, 6.74) 0.00045 | 1.82 (0.663, 4.99) 0.245 | 9.42 (3.14, 28.3) 6.31e-05 |
| Atrial fibrillation (427.21) | circulatory system | 50/331/4 | 3.49 (1.76, 6.93) 0.000351 | 1.9 (0.689, 5.23) 0.215 | 9.6 (3.2, 28.8) 5.56e-05 |
| Cardiac arrest and ventricular fibrillation (427.4) | circulatory system | 38/NA/33 | 13.6 (3.41, 54.5) 0.00022 |  | 16.1 (2.67, 96.5) 0.0024 |
| Tachycardia NOS (427.7) | circulatory system | 65/63/44 | 3.17 (1.64, 6.12) 0.000585 | 3.46 (1.37, 8.73) 0.0086 | 5.65 (1.76, 18.1) 0.00355 |
| Congestive heart failure; nonhypertensive (428) | circulatory system | 23/146/27 | 3.62 (1.95, 6.71) 4.37e-05 | 2.5 (0.967, 6.46) 0.0587 | 4.25 (1.71, 10.5) 0.00182 |
| Congestive heart failure (CHF) NOS (428.1) | circulatory system | 59/184/50 | 3.23 (1.68, 6.23) 0.000456 | 2.36 (0.889, 6.29) 0.0846 | 3.97 (1.5, 10.5) 0.00557 |
| Heart failure NOS (428.2) | circulatory system | 35/294/36 | 3.37 (1.78, 6.39) 0.000192 | 2.01 (0.738, 5.5) 0.172 | 4.24 (1.66, 10.8) 0.00252 |
| Heart failure with reduced EF [Systolic or combined heart failure] (428.3) | circulatory system | 48/102/79 | 3.93 (1.86, 8.31) 0.000346 | 3.33 (1.12, 9.86) 0.0299 | 4.04 (1.26, 12.9) 0.0187 |
| Ill-defined descriptions and complications of heart disease (429) | circulatory system | 44/97/207 | 2.71 (1.58, 4.65) 0.000283 | 2.52 (1.12, 5.7) 0.0262 | 1.84 (0.766, 4.42) 0.173 |
| Peripheral vascular disease (443) | circulatory system | 67/42/394 | 3.33 (1.66, 6.68) 0.00071 | 3.62 (1.53, 8.57) 0.00342 | 1.77 (0.374, 8.4) 0.471 |
| Other specified peripheral vascular diseases (443.8) | circulatory system | 41/40/301 | 3.9 (1.87, 8.13) 0.000275 | 3.92 (1.58, 9.74) 0.00324 | 2.25 (0.456, 11.1) 0.319 |
| Peripheral vascular disease, unspecified (443.9) | circulatory system | 29/33/332 | 4.19 (1.98, 8.85) 0.000176 | 4.12 (1.64, 10.3) 0.00251 | 2.13 (0.426, 10.7) 0.357 |
| Noninfectious disorders of lymphatic channels (450) | circulatory system | 184/47/162 | 2.92 (1.18, 7.26) 0.0209 | 6.81 (1.81, 25.6) 0.00448 | 2.89 (0.769, 10.9) 0.116 |
| Hypotension (458) | circulatory system | 42/219/28 | 2.99 (1.66, 5.41) 0.000279 | 2.07 (0.844, 5.06) 0.112 | 4.83 (1.79, 13) 0.00186 |
| Hypotension NOS (458.9) | circulatory system | 58/227/35 | 3.03 (1.63, 5.64) 0.000454 | 2.1 (0.83, 5.32) 0.117 | 5.34 (1.8, 15.8) 0.00251 |
| Pulmonary congestion and hypostasis (503) | respiratory | 145/860/13 | 4.36 (1.51, 12.6) 0.00661 | 1.15 (0.174, 7.57) 0.887 | 15.3 (3.22, 72.4) 0.000594 |
| Other pulmonary inflamation or edema (505) | respiratory | 19/23/23 | 6.63 (2.81, 15.7) 1.61e-05 | 7.26 (2.15, 24.6) 0.00144 | 9.08 (2.34, 35.2) 0.00141 |
| Empyema and pneumothorax (506) | respiratory | 30/781/2 | 4.14 (1.97, 8.7) 0.000178 | 0.794 (0.131, 4.82) 0.802 | 10.8 (3.47, 33.6) 3.99e-05 |
| Pleurisy; pleural effusion (507) | respiratory | 56/186/34 | 3.74 (1.79, 7.81) 0.000437 | 2.65 (0.873, 8.02) 0.0855 | 6.09 (1.9, 19.6) 0.0024 |
| Respiratory failure, insufficiency, arrest (509) | respiratory | 9/115/9 | 4.07 (2.36, 7.02) 4.76e-07 | 2.47 (1.04, 5.85) 0.0403 | 5.67 (2.27, 14.2) 0.000204 |
| Respiratory failure (509.1) | respiratory | 17/260/14 | 4 (2.19, 7.31) 6.53e-06 | 2.09 (0.781, 5.57) 0.143 | 5.52 (2.06, 14.8) 0.000673 |
| Respiratory insufficiency (509.2) | respiratory | 5/65/17 | 6.02 (3.2, 11.3) 2.65e-08 | 3.83 (1.4, 10.5) 0.00914 | 6.5 (2.16, 19.6) 0.00088 |
| Pulmonary insufficiency or respiratory failure following trauma and surgery (509.3) | respiratory | 1/56/1 | 11 (5.31, 22.9) 1.2e-10 | 5.45 (1.59, 18.7) 0.00699 | 15.6 (4.81, 50.5) 4.7e-06 |
| Nephrotic syndrome without mention of glomerulonephritis (580.2) | genitourinary | 37/NA/NA | 17.9 (3.9, 82.5) 0.000208 |  |  |
| Nephritis and nephropathy with pathological lesion (580.32) | genitourinary | 78/44/105 | 6.3 (2.11, 18.8) 0.000963 | 9.2 (2.03, 41.8) 0.00404 | 6.43 (1.09, 38.1) 0.0403 |
| Renal failure (585) | genitourinary | 18/59/16 | 3.45 (2.01, 5.94) 7.55e-06 | 3 (1.33, 6.74) 0.00799 | 5.54 (2.02, 15.1) 0.000855 |
| Acute renal failure (585.1) | genitourinary | 12/24/29 | 4.24 (2.37, 7.59) 1.16e-06 | 3.86 (1.64, 9.05) 0.00192 | 5.4 (1.87, 15.6) 0.00186 |
| Renal failure NOS (585.2) | genitourinary | 97/32/102 | 5.02 (1.82, 13.9) 0.00181 | 11.3 (2.38, 53.8) 0.0023 | 5.41 (1.09, 26.8) 0.039 |
| Chronic renal failure [CKD] (585.3) | genitourinary | 22/72/24 | 3.48 (1.93, 6.29) 3.56e-05 | 2.9 (1.23, 6.79) 0.0146 | 5.48 (1.91, 15.7) 0.00153 |
| Renal dialysis (585.31) | genitourinary | 34/150/78 | 6.38 (2.41, 16.9) 0.000191 | 4.92 (0.929, 26) 0.061 | 5.97 (1.37, 26) 0.0171 |
| End stage renal disease (585.32) | genitourinary | 31/48/51 | 5.54 (2.26, 13.6) 0.000179 | 7.63 (1.87, 31.1) 0.00462 | 6.22 (1.68, 23.1) 0.00632 |
| Chronic Kidney Disease, Stage IV (585.34) | genitourinary | 93/27/114 | 4.28 (1.73, 10.6) 0.00164 | 6.96 (2.03, 23.9) 0.00204 | 4.14 (0.981, 17.5) 0.0532 |
| Chronic kidney disease, Stage I or II (585.4) | genitourinary | 120/325/46 | 3.6 (1.53, 8.47) 0.00331 | 2.35 (0.617, 8.96) 0.21 | 6.36 (1.79, 22.6) 0.00419 |
| Kidney replaced by transpant (587) | genitourinary | 87/46/167 | 5.93 (1.99, 17.7) 0.00141 | 10.1 (2.05, 49.9) 0.00445 | 3.42 (0.727, 16.1) 0.12 |
| Disorders resulting from impaired renal function (588) | genitourinary | 60/31/73 | 5.03 (2.03, 12.5) 0.000487 | 8.08 (2.12, 30.7) 0.00217 | 5.03 (1.38, 18.3) 0.0143 |
| Cellulitis and abscess of arm/hand (681.3) | dermatologic | 122/22/243 | 2.87 (1.41, 5.82) 0.00352 | 4.74 (1.83, 12.3) 0.0014 | 2.12 (0.613, 7.35) 0.235 |
| Cellulitis and abscess of leg, except foot (681.5) | dermatologic | 73/17/176 | 3.18 (1.62, 6.28) 0.000822 | 4.62 (1.85, 11.6) 0.00106 | 2.4 (0.764, 7.54) 0.134 |
| Cellulitis and abscess of foot, toe (681.6) | dermatologic | 33/20/135 | 3.99 (1.93, 8.26) 0.000186 | 4.59 (1.82, 11.5) 0.00123 | 3.36 (0.875, 12.9) 0.0774 |
| Changes in skin texture (687.3) | dermatologic | 121/43/NA | 7.46 (1.94, 28.7) 0.00348 | 10.2 (2.1, 49.6) 0.00403 |  |
| Disorder of skin and subcutaneous tissue NOS (689) | dermatologic | 319/594/38 | 1.62 (0.898, 2.93) 0.109 | 1.28 (0.582, 2.82) 0.539 | 4.58 (1.7, 12.4) 0.00267 |
| Unspecified erythematous condition (695.9) | dermatologic | 49/10/NA | 8.22 (2.59, 26.1) 0.00035 | 8.62 (2.5, 29.7) 0.00064 |  |
| Osteomyelitis, periostitis, and other infections involving bone (710) | musculoskeletal | 152/3/NA | 4.65 (1.45, 14.9) 0.00957 | 20.6 (4.25, 99.9) 0.000172 |  |
| Osteomyelitis (710.1) | musculoskeletal | 142/4/NA | 5.29 (1.61, 17.4) 0.00601 | 20.6 (4.25, 99.9) 0.000172 |  |
| Acute osteomyelitis (710.11) | musculoskeletal | 118/7/NA | 8.52 (2.09, 34.7) 0.0028 | 24.8 (4.34, 142) 0.000309 |  |
| Unspecified osteomyelitis (710.19) | musculoskeletal | 143/5/NA | 5.29 (1.61, 17.4) 0.00601 | 20.6 (4.25, 99.9) 0.000172 |  |
| Bursitis (726.3) | musculoskeletal | 300/30/431 | 2.26 (0.869, 5.88) 0.0944 | 6.11 (1.92, 19.4) 0.00214 | 0.381 (0.0179, 8.12) 0.536 |
| Musculoskeletal symptoms referable to limbs (771) | symptoms | 119/11/232 | 2.54 (1.38, 4.67) 0.0028 | 4.27 (1.84, 9.91) 0.000735 | 1.92 (0.689, 5.36) 0.212 |
| Swelling of limb (771.1) | symptoms | 141/50/137 | 2.25 (1.26, 4.01) 0.006 | 3.45 (1.45, 8.23) 0.00517 | 2.19 (0.914, 5.27) 0.0787 |
| Nonspecific findings on examination of blood (790) | symptoms | 69/37/230 | 3.22 (1.63, 6.36) 0.00075 | 4.35 (1.64, 11.6) 0.00314 | 2.01 (0.679, 5.96) 0.207 |
| Complications of cardiac/vascular device, implant, and graft (854) | injuries & poisonings | 205/39/625 | 2.77 (1.12, 6.84) 0.0276 | 5.42 (1.76, 16.7) 0.0032 | 0.859 (0.141, 5.25) 0.869 |
| Complication due to other implant and internal device (859) | injuries & poisonings | 148/29/283 | 3.94 (1.45, 10.7) 0.00711 | 9.67 (2.27, 41.1) 0.00212 | 2.31 (0.492, 10.9) 0.289 |
| Open wound of foot except toe(s) alone (871.3) | injuries & poisonings | 14/NA/NA | 19.3 (5.48, 68) 4.07e-06 |  |  |
| Hormones and synthetic substitutes causing adverse effects in therapeutic use (962.3) | injuries & poisonings | 53/14/180 | 8 (2.53, 25.3) 0.000394 | 16.4 (3.13, 85.7) 0.000923 | 3.98 (0.642, 24.6) 0.138 |
| Sepsis and SIRS (994) | injuries & poisonings | 6/76/25 | 5.21 (2.88, 9.43) 4.78e-08 | 3.31 (1.25, 8.75) 0.0159 | 4.69 (1.8, 12.2) 0.00158 |
| Sepsis (994.2) | injuries & poisonings | 8/101/22 | 5.17 (2.83, 9.41) 8.24e-08 | 3.08 (1.12, 8.41) 0.0287 | 4.86 (1.85, 12.8) 0.00135 |
| Septic shock (994.21) | injuries & poisonings | 3/51/21 | 11.8 (5.06, 27.3) 1.02e-08 | 6.56 (1.72, 25) 0.00594 | 9.6 (2.46, 37.5) 0.00114 |
